# Prevalence and drivers of malaria infections among asymptomatic individuals from selected communities in five regions of Mainland Tanzania with varying transmission intensities

**DOI:** 10.1101/2024.06.05.24308481

**Authors:** Gervas A. Chacha, Filbert Francis, Salehe S. Mandai, Misago D. Seth, Rashid A. Madebe, Daniel P. Challe, Daniel A. Petro, Dativa Pereus, Ramadhani Moshi, Rule Budodo, Angelina J. Kisambale, Ruth B. Mbwambo, Catherine Bakari, Sijenunu Aaron, Daniel Mbwambo, Samuel Lazaro, Celine I. Mandara, Deus S. Ishengoma

## Abstract

**Background:** Malaria is still a leading public health problem in Tanzania despite the implementation of effective interventions for the past two decades. Currently, the country experiences heterogeneous transmission and a higher malaria burden in some vulnerable groups, threatening the prospects for elimination by 2030. This study assessed the prevalence and drivers of malaria infections among asymptomatic individuals in selected communities from five districts within five regions with varying endemicity in Mainland Tanzania.

**Methods:** A community cross-sectional survey was conducted in selected communities (covering 15 villages) from five districts, one each from five regions of Kagera, Kigoma, Njombe, Ruvuma, and Tanga from July to August 2023. Asymptomatic participants aged ≥6 months were recruited and tested with rapid diagnostic tests (RDTs) to detect malaria parasites. Demographic, anthropometric, clinical, parasitological, housing type, and socio-economic status (SES) data were captured using questionnaires configured and installed on Open Data Kit (ODK) software run on tablets. The association between parasite prevalence and potential drivers of malaria infections among asymptomatic individuals were determined by univariate and multivariate logistic regression, and the results were presented as crude (cOR) and adjusted odds ratios (aOR), with 95% confidence intervals (CI).

**Results:** Testing involved 10,228 individuals and 3,515 (34.4%) had RDT positive results. The prevalence varied from 21.6% in Tanga to 44.4% in Kagera, and ranged from 14.4% to 68.5% in the different villages, with significant differences among regions and villages (p<0.001). The prevalence and odds of malaria infections were significantly higher in males (aOR =1.32, 95% CI:1.19 -1.48, p<0.01), under-fives (aOR = 2.02, 95% CI: 1.74 - 2.40, p<0.01), school children [aged 5 – <10 years (aOR =3.23 95% CI: 1.19–1.48, p<0.01) and 10–15 years (aOR = 3.53, 95% CI: 3.03 – 4.11, p<0.01)], and among individuals who were not using bed nets (aOR = 1.49, 95% CI: 1.29 –1.72, p<0.01). The odds of malaria infections were also higher in individuals from households with low SES (aOR = 1.40, 95% CI:1.16 – 1.69, p<0.001), living in houses with open windows (aOR = 1.24, 95% CI: 1.06 – 1.45, p<0.01) and holes on the wall (aOR = 1.43, 95%CI 1.14 – 1.81, p<0.01).

**Conclusion:** There was a high and varying prevalence of malaria infections in the surveyed regions/villages. The odds of malaria infections were higher in males, school children, individuals who did not use bed nets, and participants with low SES or living in poorly constructed houses (with open windows and holes on walls). These findings provide useful information for identifying high-priority vulnerable groups and areas for implementing targeted malaria control interventions for reducing the burden of asymptomatic infections.

## Background

Tanzania has experienced a significant decline of malaria burden in the past two decades [1]. This has been achieved after adopting and implementing the recommended World Health Organization (WHO) measures for enhanced malaria control and elimination [2,3]. Despite a significant decline, malaria is still a leading public health problem in Tanzania and the country experiences heterogeneous transmission at macro and microgeographic levels [2,4]. According to the WHO World Malaria report of 2023, there were an estimated 249 million malaria cases and 608,000 deaths globally in 2022 and majority of these were from the WHO region for Africa (WHO - Afro) [5]. Tanzania was among the eleven countries with the highest malaria burden accounting for over 4.4% of all malaria deaths globally in that year [5]. Over 93.0% of the Tanzanian population lives in areas where transmission occurs, with the entire population living in areas with ongoing transmission and at risk of malaria infections [6]. The country experiences varying levels of malaria burden ranging from very high and stable to very low transmission intensities [7,8]. *Plasmodium falciparum* is the leading cause of malaria in Tanzania contributing to 96% of malaria cases, while a few cases (4.0%) are due to other *Plasmodium* species including *P. ovale spp*, *P. malariae* and *P. vivax* [9,10].

Despite the progress made, Tanzania still faces contemporary challenges that may potentially limit its progress toward malaria elimination. In recent years, there have been reports of resistance to insecticides by mosquito vectors [11–13] and parasites to widely used antimalarial drugs [14,15], including the emergence of artemisinin partial resistance (ART-R) in Kagera region [16,17]. In addition, recent studies reported the presence of parasites with histidine-rich protein 2/3 (*hrp2/3)* gene deletions that affect the performance of histidine-rich protein 2/3 (HRP2/3) - based rapid diagnostic tests (RDTs) but at low prevalence [18,19]. There is also a high risk of emergence and spread of an invasive *Anopheles stephensi* vector that has been reported in Kenya and other countries in the Horn of Africa particularly in Ethiopia [20–22]. These reports suggest that the country needs to intensify surveillance to detect emergence of new threats, track and control the already reported threats and ensure progress to the elimination targets is not impacted.

Based on the WHO recommendations, the Tanzania National Malaria Control Programme (NMCP) has been implementing most of the key effective interventions for vector control and case management together with other initiatives such as preventive therapies. For vector control, the interventions used by NMCP in Tanzania include insecticide treated bed nets (ITNs), indoor residual spraying (IRS), and larval source management (LSM) using effective larvicides [2]. Case management interventions primarily focus on timely and accurate parasitological diagnosis and confirmation with RDTs and effective malaria treatment using artemisinin-based combination therapy (ACT) [3,6]. Although WHO recommends several preventive therapies [23], Tanzania is currently using intermittent preventive treatment for pregnant women (IPTp) using sulfadoxine-pyrimethamine (SP), targeting to deliver two or more doses to all pregnant women from their first trimester [24]. Plans are also underway to deploy and implement intermittent preventive treatment to school children and infants (also known as perennial malaria chemoprevention). Thus, strategies are urgently needed to support the ongoing control efforts and enhance progress toward the 2030 malaria elimination targets.

Due to scaled up interventions in Tanzania, malaria burden has significantly declined with the overall prevalence dropping from 18% in 2008 to 8% in 2022 [1]. The transmission of malaria is currently heterogeneous with a large proportion of the regions (about 38%) located in areas with low to very low transmission intensities (covering the central corridor, northern and south-western parts), sand-witched by moderate transmission areas on both sides and high malaria burden in north-western, north-eastern, southern and western regions [2,4,25] (**Figure 1**). Hence, novel interventions are urgently needed to support the NMCP efforts to get back on track and progress towards the national elimination targets through deployment and use of new and current interventions based on the local burden and transmission intensities [3].

**Figure 1:**
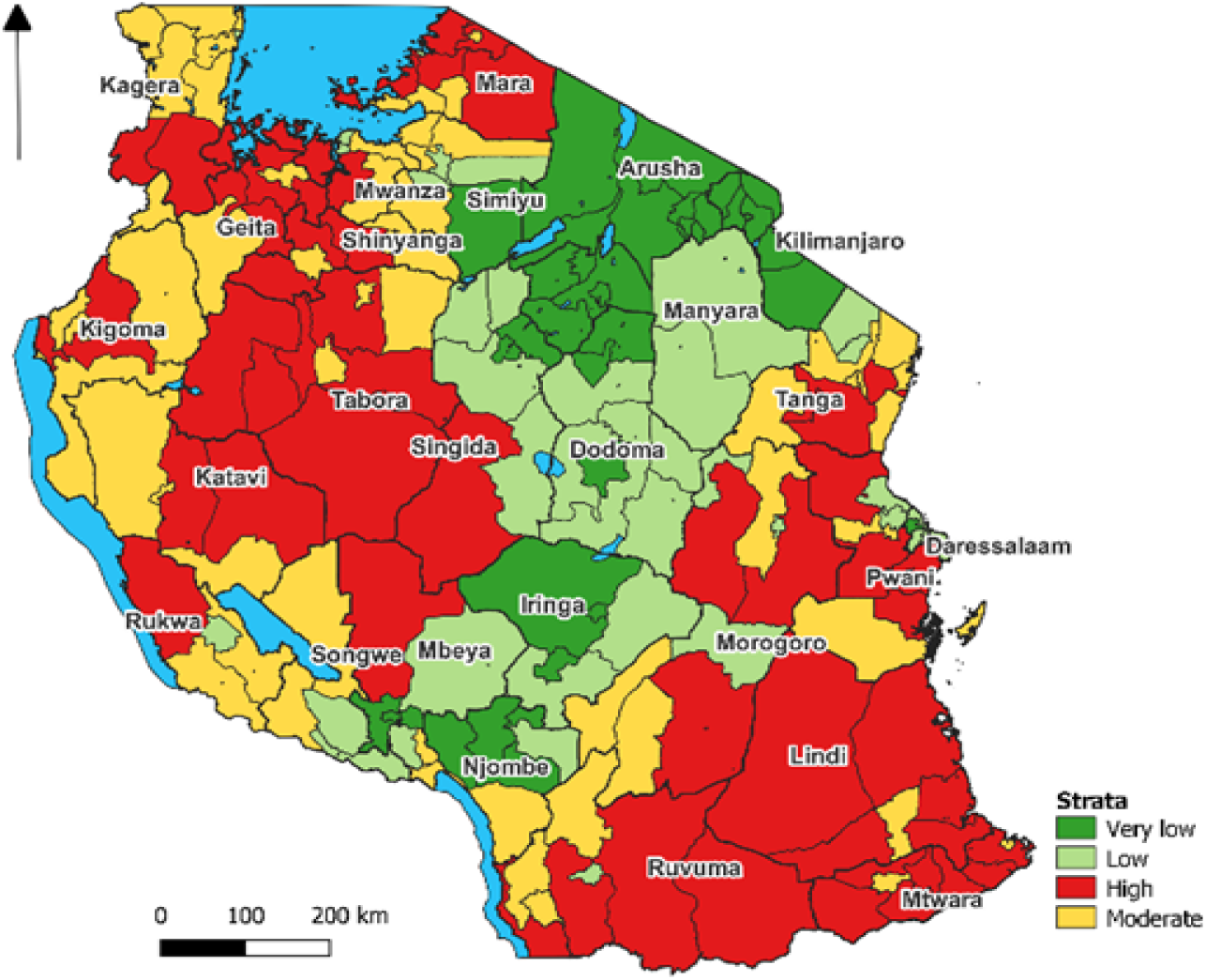
Map of Tanzania showing malaria burden microstratification by council level, 2022 [28].

In Tanzania, extensive research, surveillance, and control efforts have been undertaken over the past two decades [25] and have contributed to increased awareness, improved diagnostics, and more effective treatment strategies for malaria [26]. However, amidst these commendable efforts, relatively little attention has been given to asymptomatic individuals, leaving a significant gap in our understanding of their prevalence and contribution to malaria transmission in the communities. Asymptomatic individuals serve as reservoirs of infections and contribute to the persistence of transmission impending malaria reduction and elimination efforts [27]. The scarcity of data on the scope and prevalence of asymptomatic malaria presents a critical gap in malaria control efforts. To address the gap, this study aimed to assess the prevalence and drivers of malaria infections among asymptomatic individuals through cross-sectional community survey (CSS) in five regions of Mainland Tanzania with varying transmission intensities. The findings from this study help NMCP and other stakeholders enhance the current surveillance system and close the gaps in the ongoing elimination efforts through targeting of areas with high prevalence of malaria infections and vulnerable groups.

## Methods

### Study design and sites

This was a community-based CSS which was conducted in five regions of Mainland Tanzania from July to August 2023. It was implemented as one of the components of the main project on molecular surveillance of malaria in Tanzania (MSMT). The MSMT project was implemented in 13 regions of Mainland Tanzania in 2021 and 2022, and was later extended to cover all 26 regions from 2023 (Ishengoma et al, unpublished data). The five regions covered by this study have different malaria transmission intensities and they include three regions from high (Kagera, Kigoma and Ruvuma), one from each moderate (Tanga) and low (Njombe) transmission intensity (**Figure 2**). In Kagera region, the survey covered five villages (Kitoma, Kitwechenkura, Nyakabwera, Rubuga, and Ruko) from Kyerwa district as recently described [29]. These five villages in Kyerwa district are located in an area where recent reports showed the emergence of parasites with ART-R [16,17]. In Kigoma, the study was conducted in two villages of Nyankoronko and Kigege (in Buhigwe district) and in Ruvuma, the surveys were done in four villages (Chiulu, Lipingu, Lundo, and Ngindo) in Nyasa district as described in previous publications [18,30,31]. In Njombe region, the surveys were done in Kipangala, which is a new village under the MSMT longitudinal surveillance component aiming at monitoring the trend and pattern of parasite populations (Ishengoma D unpublished data) and supporting capturing and tracking parasites with *hrp2/3* gene deletions which were recently detected in the area, albeit at low prevalence [18]. In Tanga region, the survey was done in three villages of Magoda, Mamboleo, and Mpapayu, in Muheza district where different surveys were done from 1992 and the community has been under surveillance ever since as reported elsewhere [32–34]. Individuals living in these communities are served by dispensaries that have been participating in the longitudinal component of the MSMT that was initiated in July 2022 (in Kigoma, Ruvuma and Tanga regions) and March 2023 (in Kagera and Njombe regions) to longitudinally undertake genomic studies of malaria parasites as described elsewhere (Kanyankole G. et al., unpublished data).

**Figure 2:**
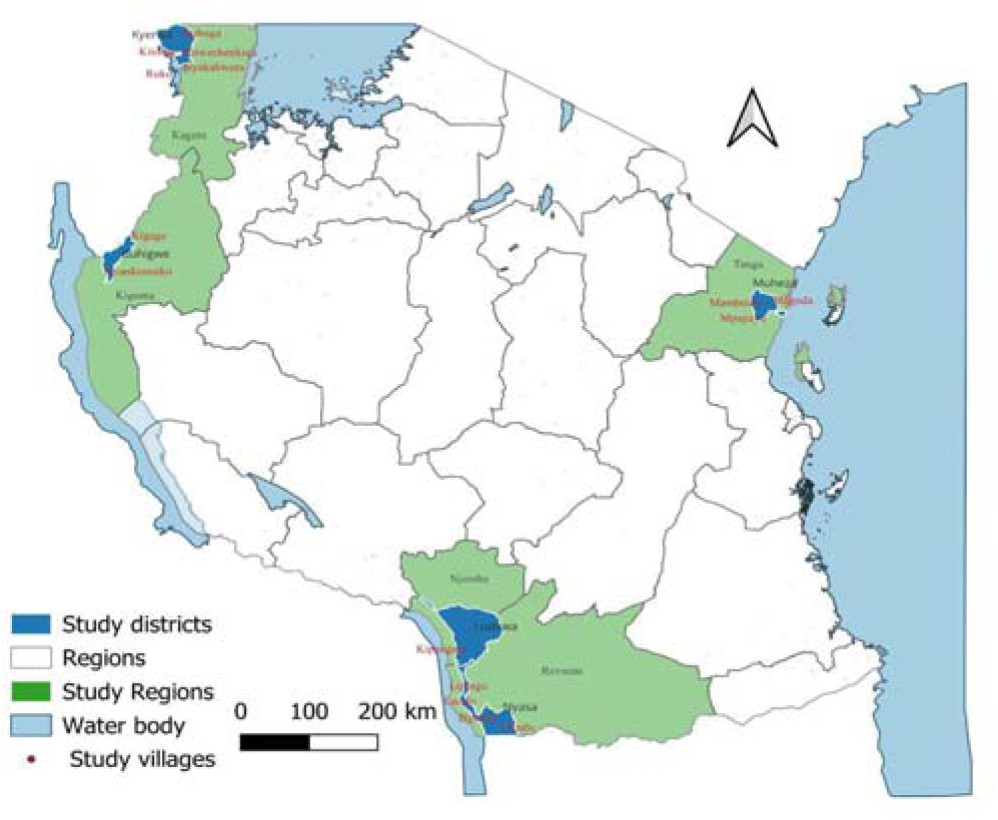
Map of Tanzania showing the regions, districts and study villages.

### Study population and recruitment of participants

The study was conducted within the CSS as described earlier [29] and targeted about 30% of all individuals aged ≥6 months living in the study villages who were registered during previous census surveys undertaken by the MSMT project before the CSS. Briefly, inclusion criteria for the CSS included age ≥6 months, residence in one of the study villages and providing an informed consent. The study did not include individuals from villages not covered by the MSMT project or those who declined to give an informed consent. All community members were informed about the study by the community/village/hamlet sensitization teams, which passed the information to the community using a loudspeaker for 1 - 2 days before the CSS. Each study village has 3 - 5 hamlets and registered members from each hamlet were invited to meet the study team at the CCS post on designated survey days. On each day, 1 - 2 hamlets were invited and household members who failed to attend on their scheduled day(s) were allowed to visit the team on any other day in the same village or even in a nearby village in case the survey covered more than one village in the same area. Thus, participants were recruited conveniently based on their willingness to visit the recruitment posts and consent to take part in the CSS.

### Data collection procedures

Prior to the CSS, census surveys were conducted in each village to collect demographic data of community members, register households, and obtain socio-economic status (SES), environmental, land use and geographical positioning system (GPS) data. Members of the selected villages in each region and their households were enumerated and given unique identification numbers as described earlier [29]. During the CSS, all participants were identified using their unique IDs that were given during the census and data collection was done as previously described [29]. Briefly, each participant had their identities verified and assigned study IDs specific to the CSS and provided with registration cards. Study procedures included obtaining consent and/or assent (for participants aged 7 - 17 years), and interviews to collect socio-demographic and malaria prevention information. Participants were directed to the next station for anthropometric measurements (body weight, height and axillary temperature) and then to the laboratory where finger pricking was done for the detection of malaria parasites using RDTs and collection of dried blood spots (DBS) on filter papers (Whatman No. 3, GE Healthcare Life Sciences, PA, USA). The RDTs which were used in the CSS included Abbott Bioline Malaria Ag Pf/Pan (Abbott Diagnostics Korea Inc., Gyeonggi-do, Korea) and Malaria Pf/Pan Ag Rapid Test (Zhejiang Orient Gene Biotech Co. Ltd, Zhejiang, China). From the same finger prick, DBS and blood smears (thin and thick) were also collected for further laboratory analyses (analysis is underway and data will be reported in our future reports). The final section was for clinical assessment where participants were assessed by study clinicians to obtain data on their history of illness and any treatment taken two weeks before the survey. Each participant had a physical examination and clinical diagnosis done, and those who needed treatment in case of a positive malaria test or any other illness were managed according to the national guidelines for the management of malaria [35] or the guidelines for other febrile conditions [36].

### Data management and analysis

Data collection was done using questionnaires which were developed and configured in Open Data Kit (ODK) software and installed on tablets. Subsequently, the collected data was transferred daily to the central server at the National institute for medical research (NIMR), Dar es Salaam. Data quality checks were integrated into the databases through consistency checks, where any unexpected values were flagged for rejection, prompting corrections by the field team. The data was exported to excel for additional cleaning and later transferred to STATA version 13 (STATA Corp Inc., TX, USA) for final cleaning and analysis. Initial descriptive analysis was done to provide baseline information on the study populations and their demographic characteristics. Chi-square tests were used to assess the relationship between categorical variables. The results were presented in tables, figures and texts.

Multilevel logistic regression was used to assess the association between malaria infections and other covariates such as age groups, sex, household size, education levels, and geographic locations. Variables with p<0.25 in the univariate analysis were fitted into multivariate models. Hierarchical model-building strategies were used for further analysis whereby the first model was made by adjusting for individual-level variables such as sex, age group, and the use of bed nets the night before the survey. In the second (model II), the analysis included both individual and household characteristics such as household size, household wealth index, and the type of houses inhabited (focusing on the type of windows, walls and presence/absence of eaves). Principal component analysis (PCA) was used to determine the SES of the households based on the assets owned by each family, using the data collected during the census survey to compute the wealth index of the family as previously described [29]. The variables included in PCA were household possessions and assets such as the occupation of head of households, number of rooms in the house, ownership of items such as mobile phones, motorcycles, bicycles, and domestic animals such as cattle, goats, chicken and pigs. Other assets were land owned by the family and the number of acres cultivated, the source of drinking water, lighting and cooking energy, and the type of toilet. The scores of the first component with eigenvalues >1 were used to create household wealth index/SES (categorized as high, moderate and low). Inter-cluster correlation (ICC) was used to estimate the proportion of variances contributed by household characteristics and other parameters such as the Akaike information criterion (AIC) and Log-likelihoods test (-2LL) tests were used to assess the goodness of fit. The association between variables was reported as crude (cOR) or adjusted odds ratios (aOR) and p-value ≤0.05 was considered statistically significant.

## Results

### Demographic characteristics of participants

The study covered 15 villages from the five districts of Buhigwe (Kigoma region), Kyerwa (Kagera), Ludewa (Njombe), Muheza (Tanga) and Nyasa (Ruvuma) which had a population of 40,116 individuals in 8,881 households. Of the entire population, 10,228 (25.5%) were recruited in the CSS and majority of them were females (60.3%), but their proportions varied significantly in the different districts (p=0.006). Most of the participants (47.9%) were aged ≥15 years and the overall median age was 14.1 years (IQR = 7.2 - 38.1 years). The age of participants varied significantly among the districts with the lowest median age in Buhigwe (11.9 years) and the highest was in Ludewa (17.6 years) (p<0.01). A high proportion of the participants (43.8%) had completed primary education and their main occupation was farming (42.7%). History of fever in the past 48 hours before the survey was reported by 20.6% of the participants but only 2.3% had fever at presentation with axillary temperature ≥37.5^0^C (**Table 1**).

**Table 01:**
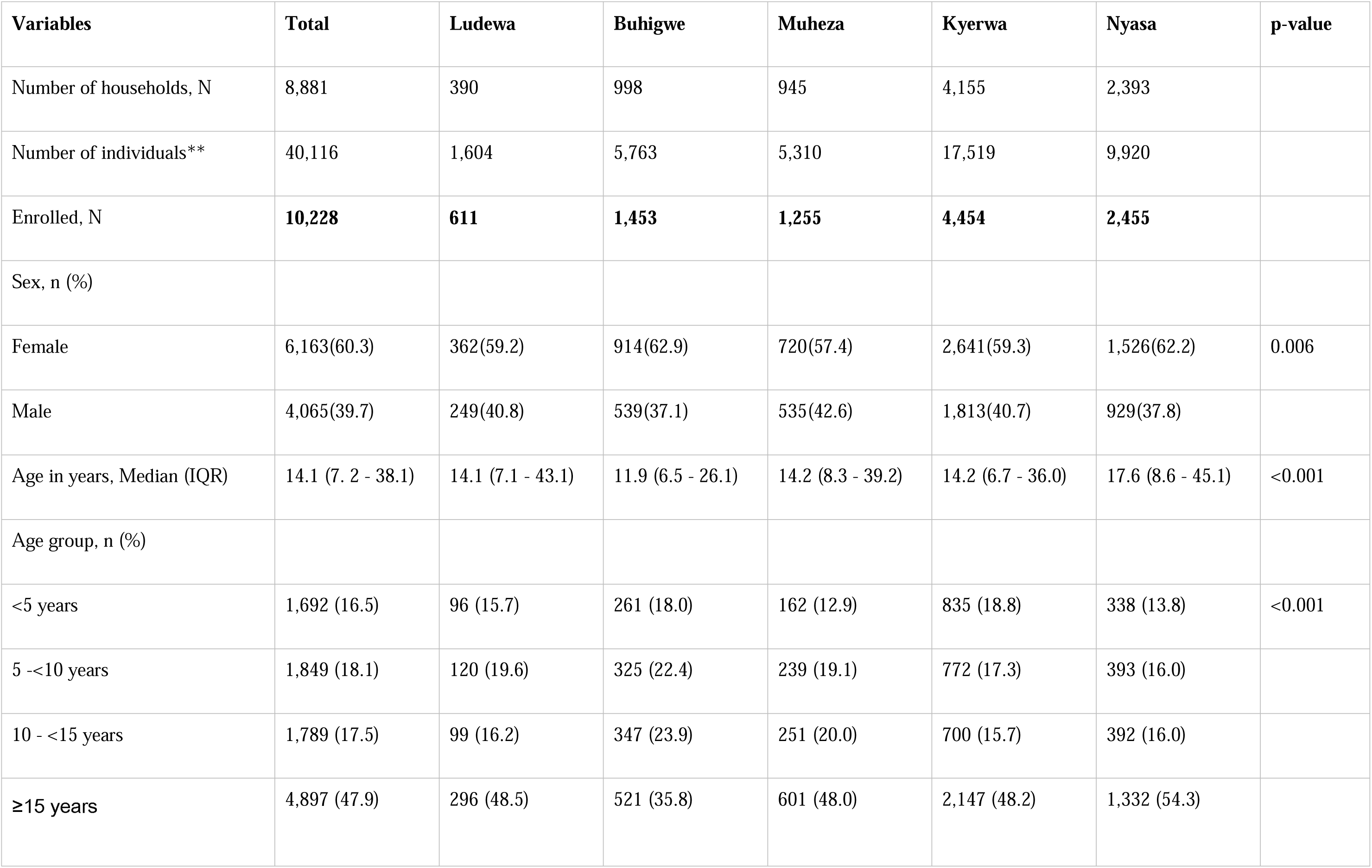
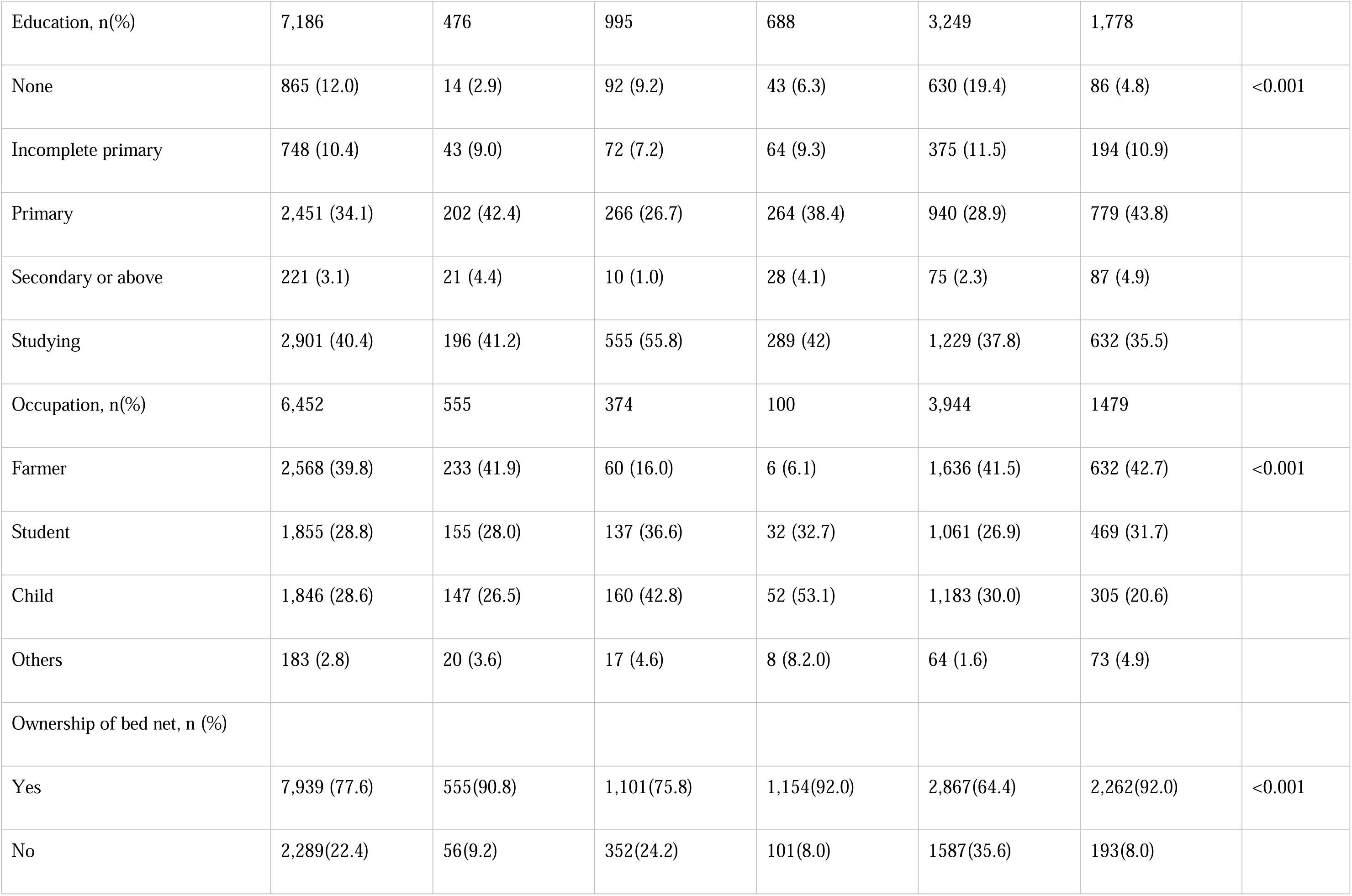
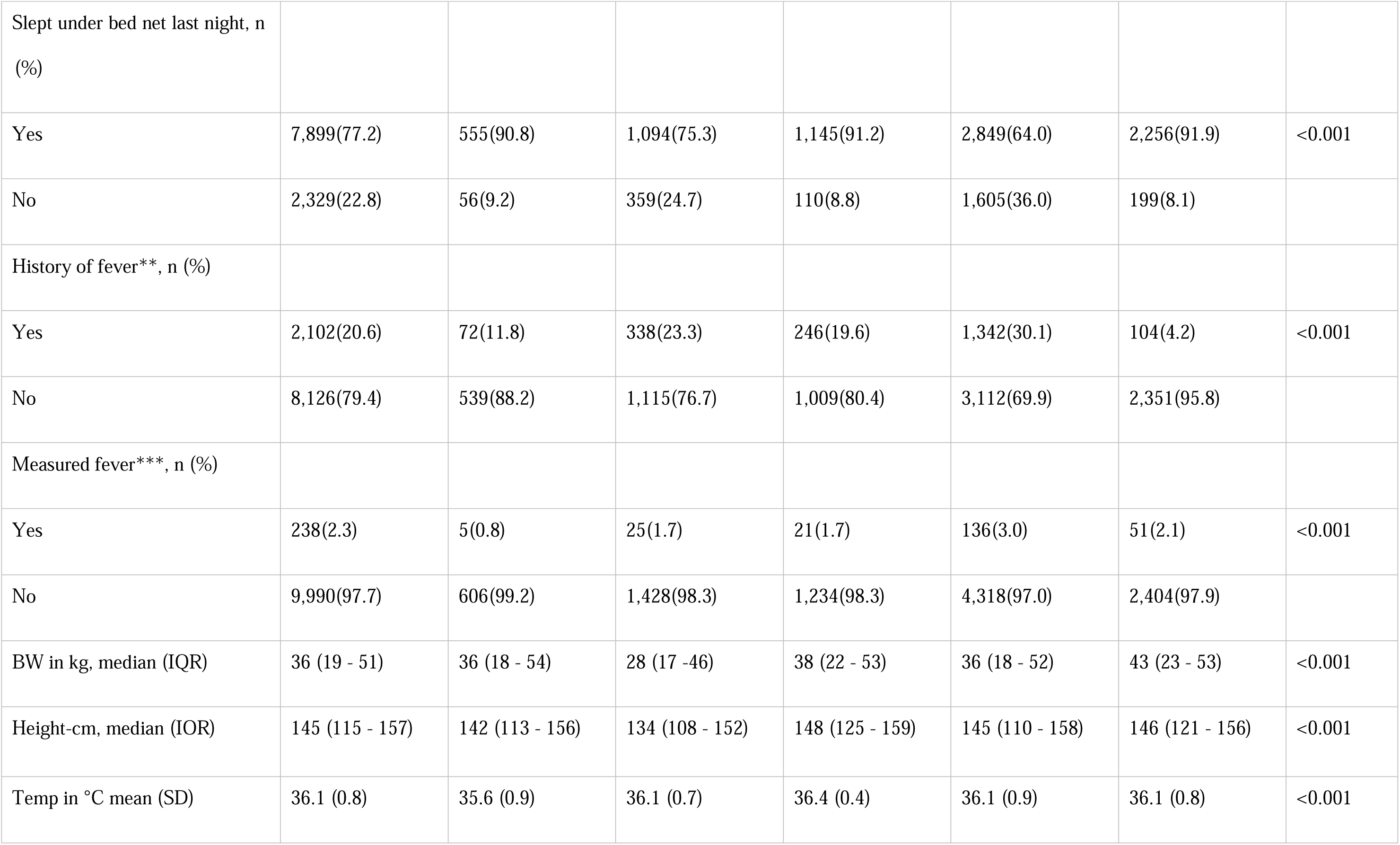
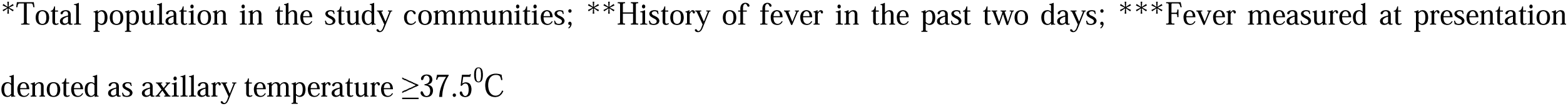
Demographic characteristics of study participants.

### Prevalence of malaria

In all districts, 34.4% (3,515/10,228) of the participants had positive results by RDTs. The lowest prevalence (21.6%) was in the three villages of Muheza (Tanga region) while the highest was in the five villages of Kyerwa district, in Kagera region (44.4%), and the differences in prevalence among the districts were statistically significant (p<0.001). The prevalence was significantly higher in males (38.9%, p <0.001), school children ((aged 5 to <10 years (44.6%, p<0.001) and those aged 10 to <15 years (47.2%, p<0.001)), participants who did not use bed nets the night before the survey (41.5%, p<0.001), and those who reported history of fever in the past two days (77.4%, p<0.001) or had fever at presentation (62.6%, p<0.001). Under-fives from all districts had lower prevalence than school children but they had higher prevalence compared to adults (≥15 years old) (**Table 2**).

**Table 02:**
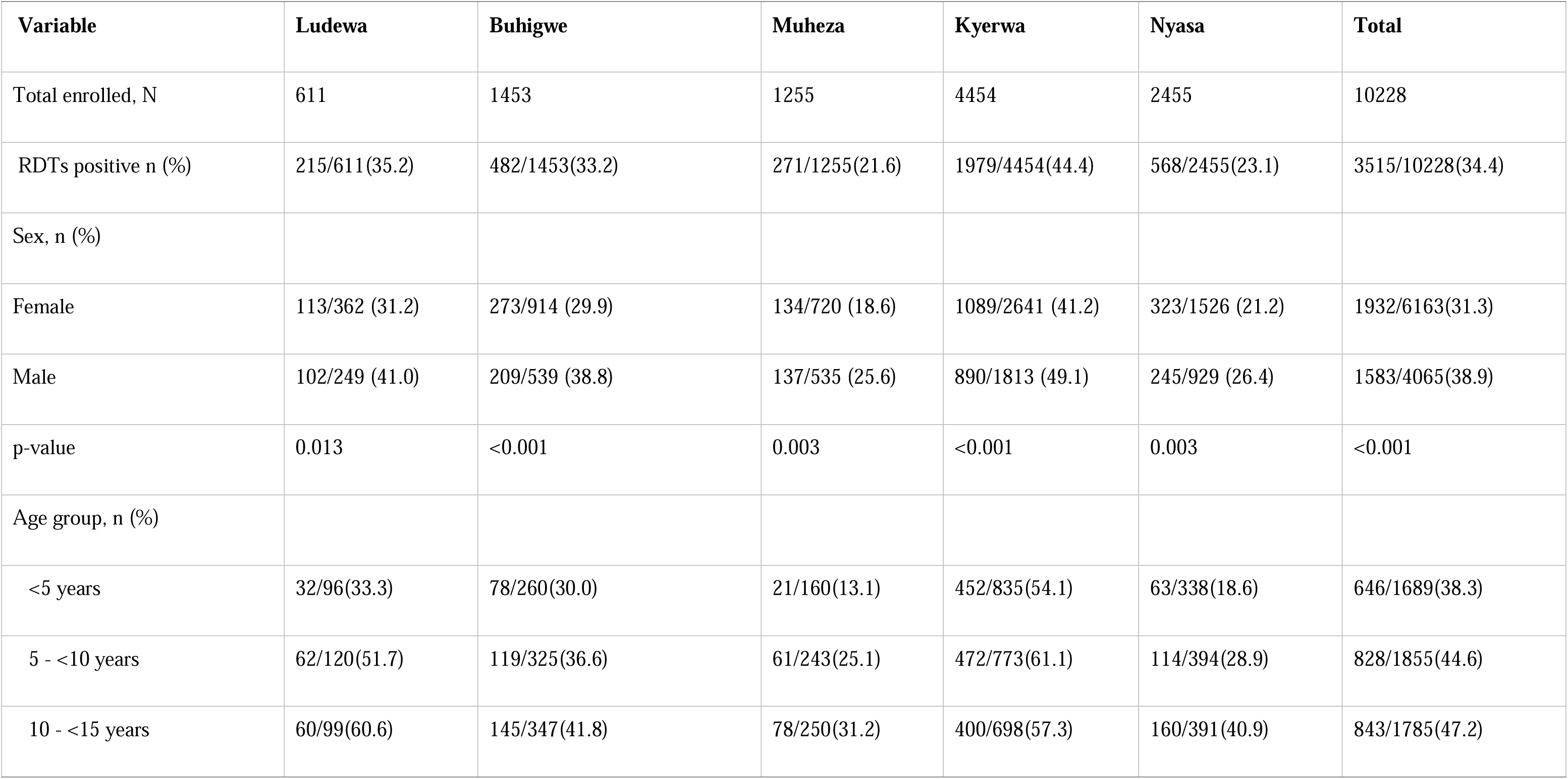
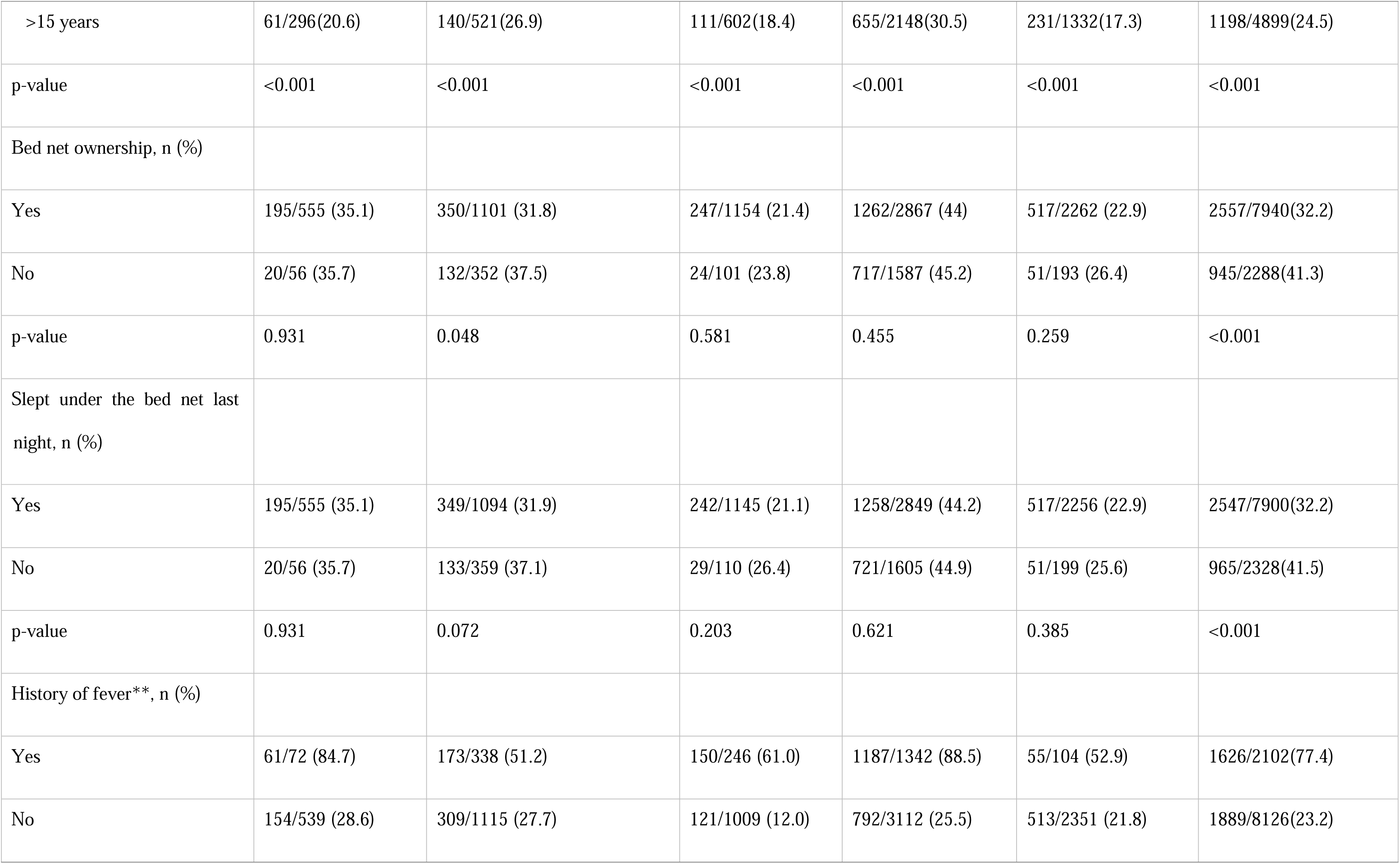
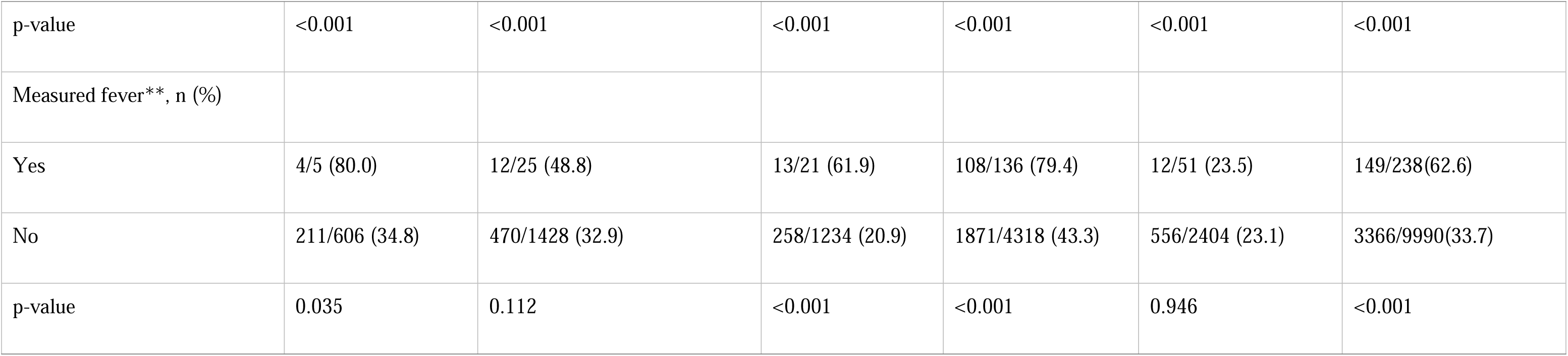
Prevalence of Malaria infections in five districts.

In the districts with more than one village, the prevalence of malaria infections varied significantly among villages (p<0.001). Overall, the villages in Kyerwa had higher prevalence compared to other districts, with the lowest of 14.5% and the highest of 68.5%. Villages in Muheza had the lowest parasite prevalence, ranging from 16.3% to 29.0%. The differences in prevalence among the villages were statistically significant in the three districts of Muheza, Kyerwa, and Nyasa (p<0.001), while no difference was observed in Buhigwe (p = 0.056). In all districts, only four villages had prevalence of less than 20%, including two in Muheza and one village each in Kyerwa and Nyasa; and the highest heterogeneity was observed in Kyerwa (**Figure 3 and Table 2**).

**Figure 3:**
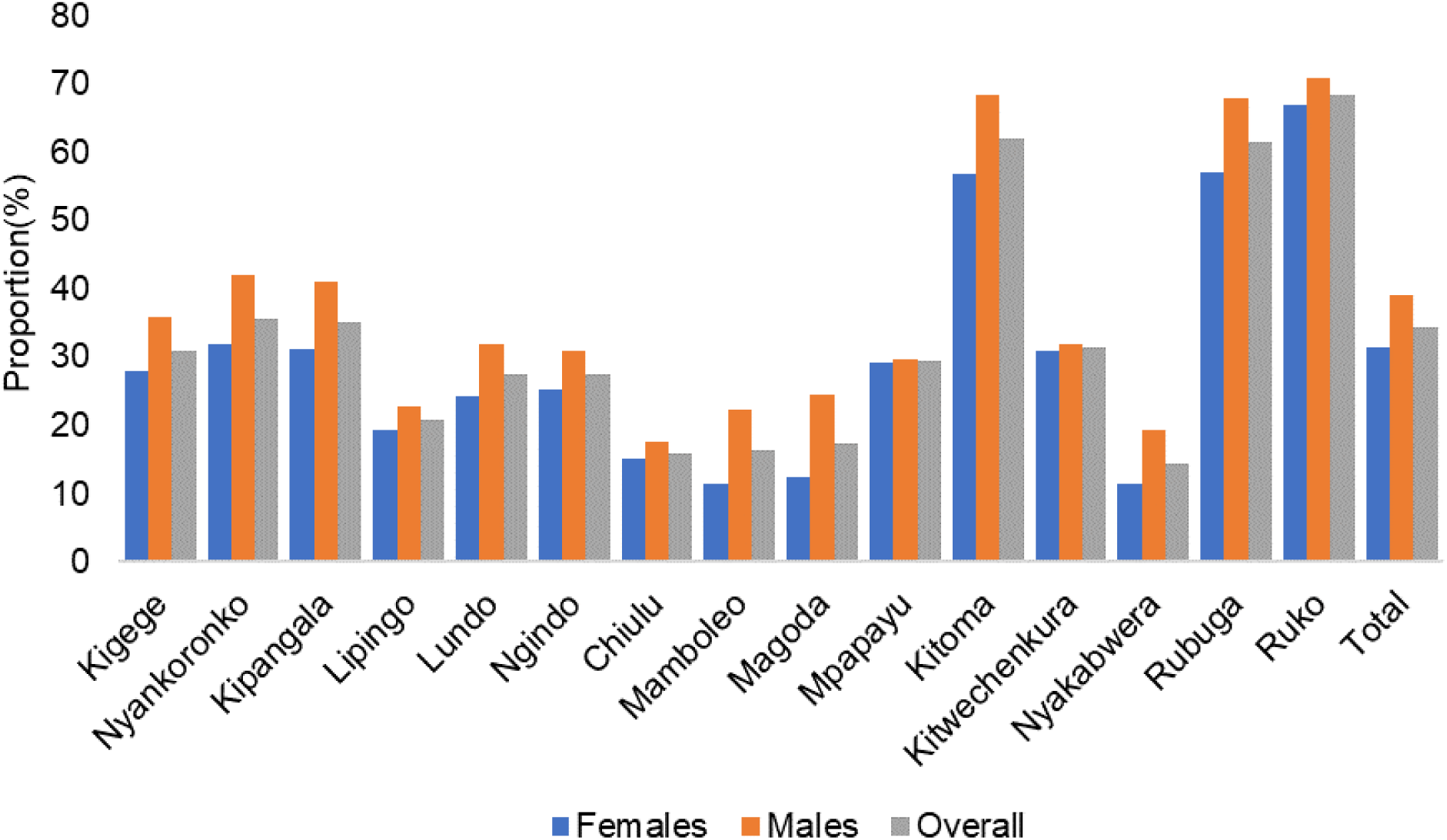
Prevalence of malaria infections by RDTs among male and female participants from different villages in five districts.

Among the different age groups, school children had the overall highest prevalence of malaria infections (45.9%), followed by under-fives (38.3%), and the lowest prevalence (24.5%) was observed in adults (**Figure 4)**. Across villages, the prevalence was higher among school children compared to other age groups. Conversely, adults in most villages had the lowest prevalence except for three villages from Muheza districts (Mamboleo, Magoda, and Mpapayu) where under five had the lowest prevalence compared to other groups. With the exception of four villages (Chiulu - Buhigwe, Mpapayu - Muheza, and Kiwechenkuara and Rubuga in Kyerwa), the highest prevalence of malaria infections was observed in older school children (aged 10 - <15 years) (**Figure 5 and Table 2**). Malaria prevalence was higher among males aged 15+ years in all villages compared to females. In under-fives, the pattern of prevalence in males and females was different in the study villages, whereby some villages had higher prevalence in males and in other villages, the prevalence was higher in females. In school children, the prevalence was higher in males but not in all villages (**Supplementary Figure S1).**

**Figure 4:**
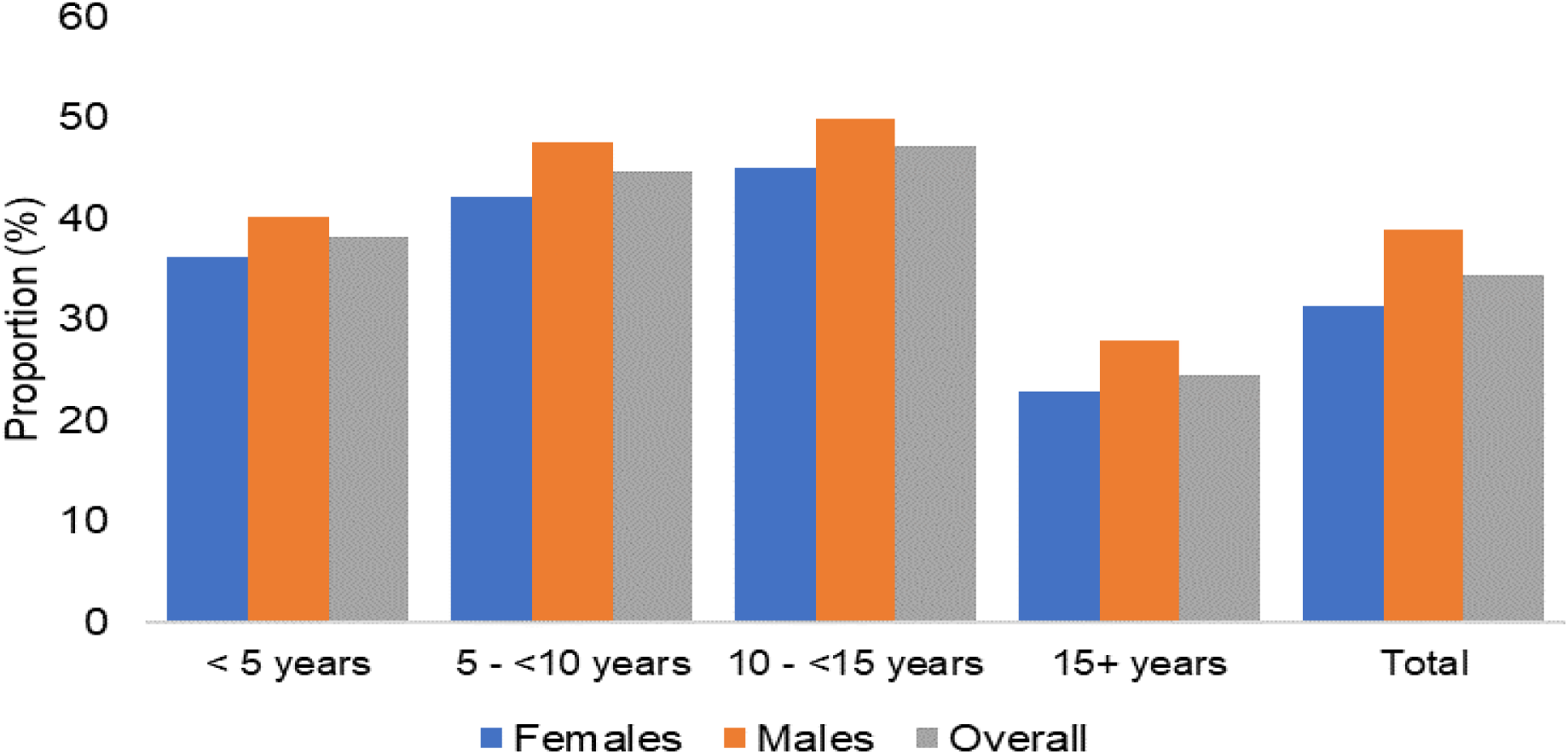
Prevalence of malaria infections by RDTs among male and female participants of different age group.

**Figure 5:**
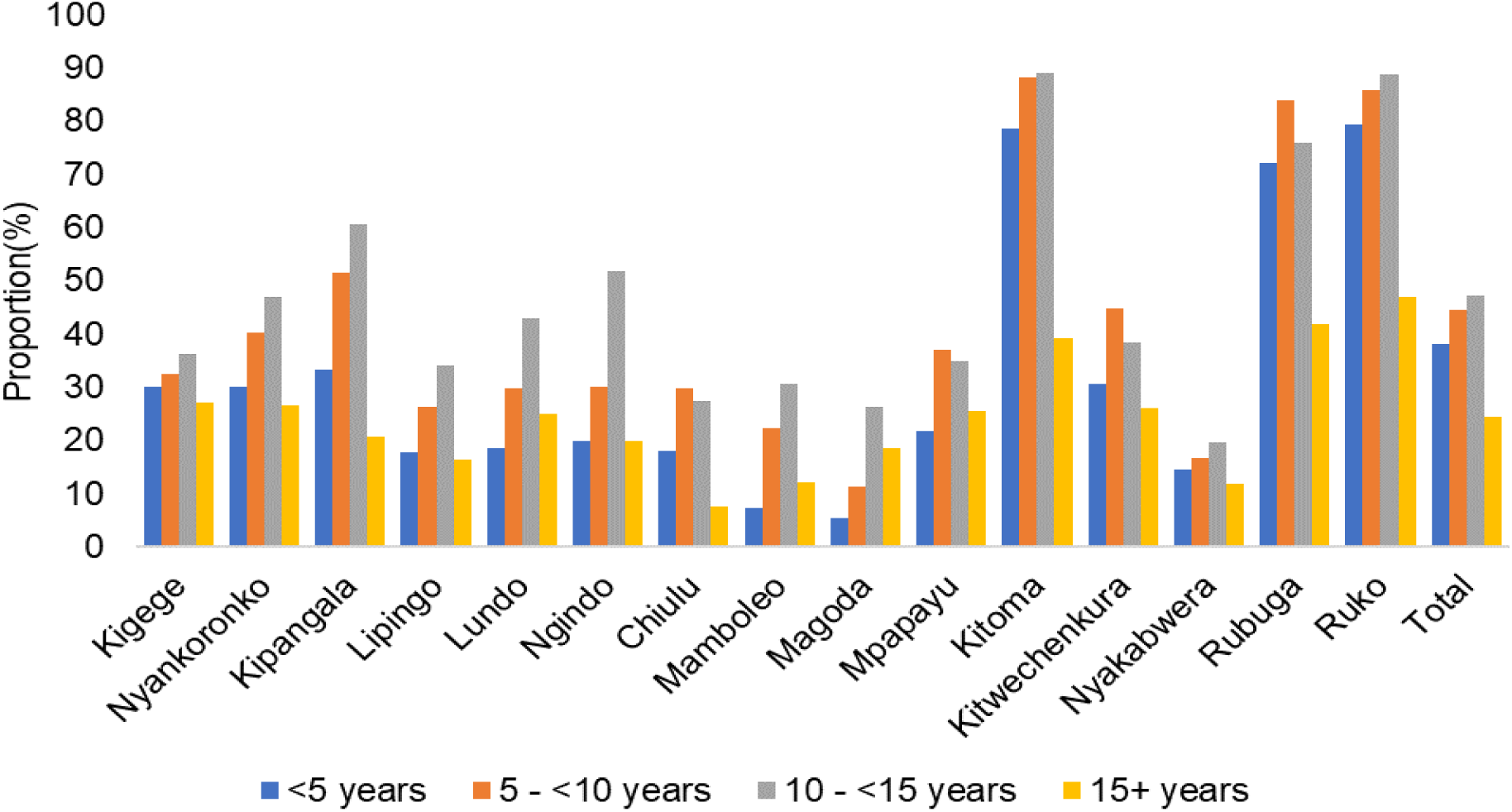
Prevalence of malaria infection among individuals of different age groups in the study village from the five districts.

### Bed net ownership and use, and prevalence of malaria infections

About 78.0% of the participants had bed nets and 77.2% slept under the nets the night before the survey (**Table 1**). Bed net ownership was significantly higher (≥91%) in three districts (Ludewa, Muheza, and Nyasa) compared to Buhigwe (75.8%) and Kyerwa (64.4%). A similar trend was observed in bed net usage with the rates exceeding 91.0% in the three districts of Ludewa, Muheza, and Nyasa and lower rates of 75.3% in Buhigwe and 64.0% in Kyerwa (p<0.001 for all comparisons). Bed net ownership and use were significantly higher among females in the districts of Kyerwa (p≥0.005) and Muheza (p<0.001) while it was similar among male and female participants in the other three districts (Buhigwe, Ludewa and Nyasa; p≥0.218) (**Figure 6).** In all age groups, under-fives had higher rates of bed net ownership across all districts except in Muheza. In contrast, school children in Ludewa and Kyerwa, as well as adults in Buhigwe and Nyasa, had comparatively lower rates of bed net ownership. In Muheza district, bed net ownership was higher among school children and lower among adults compared to others (**Figure 7A).** Similarly, bed net use was notably higher among under-fives across all districts except Muheza. Usage of nets was lower among school children in Ludewa and Kyerwa, and among adults in Buhigwe and Nyasa districts compared to other age groups. In Muheza district, bed net usage was higher in school children and lower among adults (p=0.001) (**Figure 7B).**

**Figure 6:**
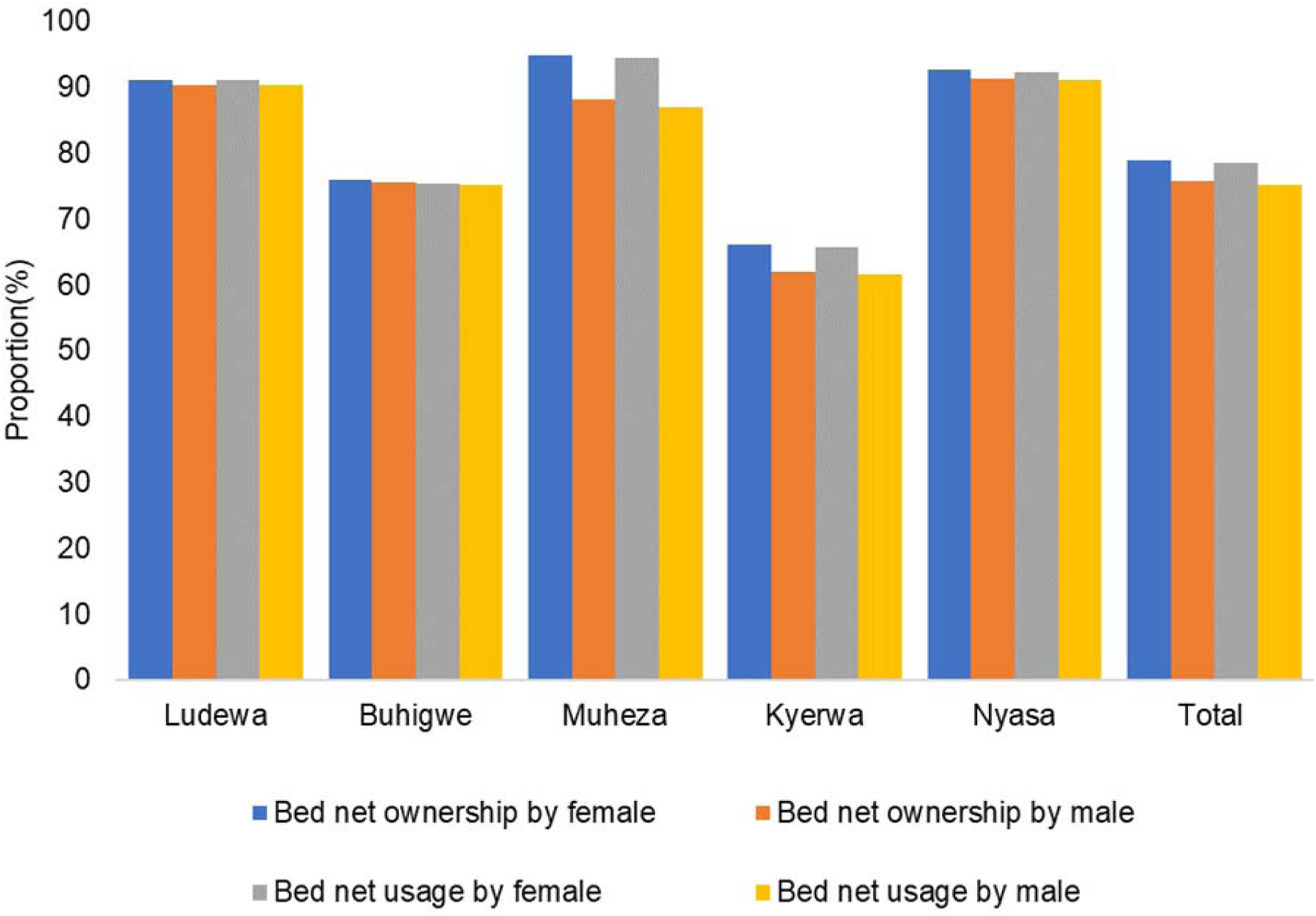
Bed net ownership by usage by sex in five districts.

**Figure 7:**
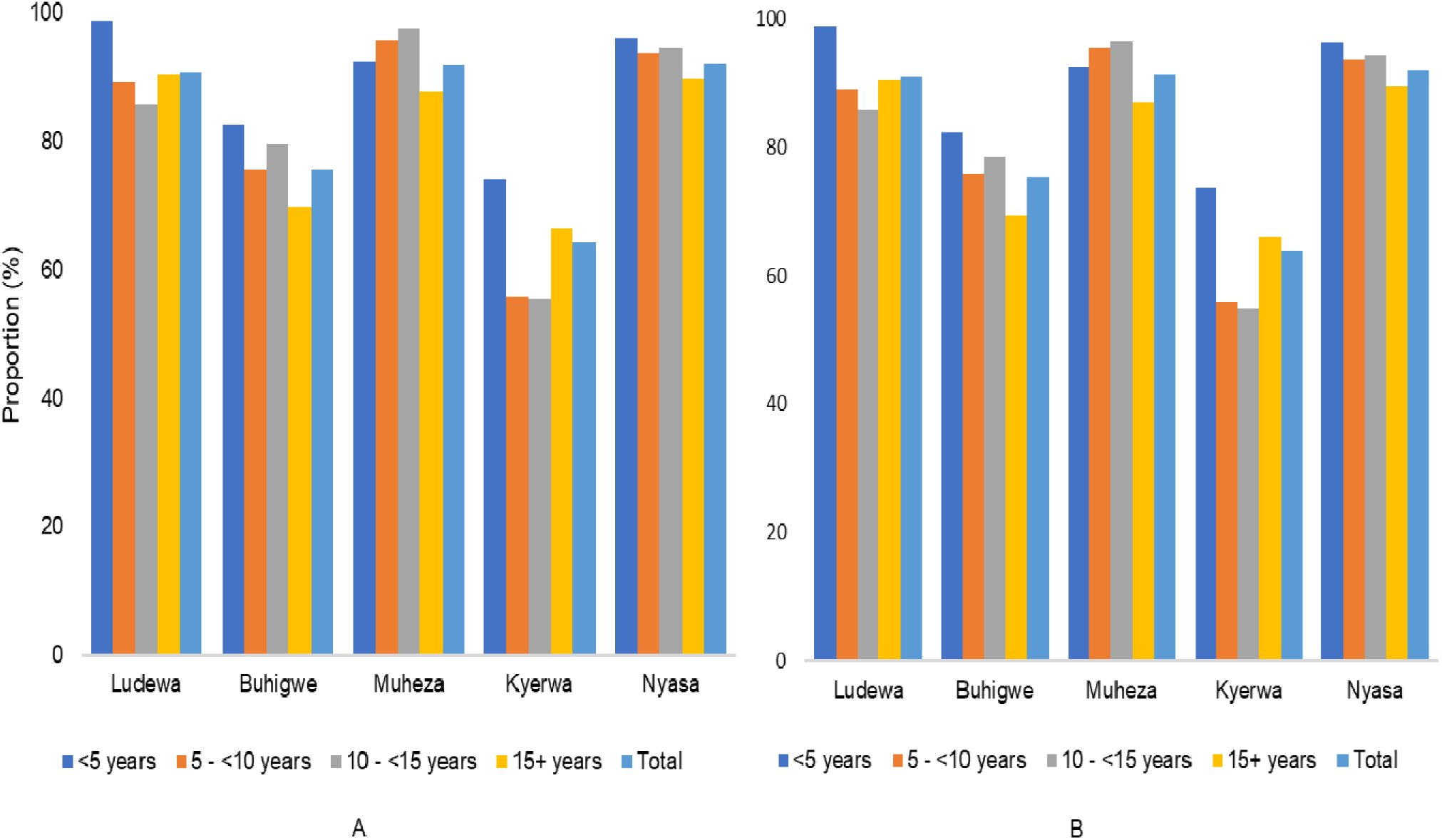
Bed net ownership (A) and use (B) among individuals of different age groups in the five districts.

A higher prevalence of malaria infection (over 41.0%) was observed among individuals who did not own or sleep under the bed net compared to their counterparts and increased rates were observed among participants from Kyerwa who neither owned (45.2%) nor slept (44.9%) under bed nets. A similar trend was observed in all other districts. Overall, there was a significant association between malaria prevalence and bed net ownership as well as sleeping under bed nets in the night before the survey (p<0.001) (**Table 2**).

### Prevalence of malaria infections and household characteristics

The prevalence of malaria infections was significantly higher in households with five or more people (35.4%) compared to those with fewer members (30.0%) (p<0.01). Similarly, individuals from households with low SES had significantly higher malaria prevalence (37.9%) compared to those with moderate (33.3%) and higher SES (31.3%) (p<0.01). The type of walls of the houses were also associated with malaria (p<0.01) whereby individuals living in houses made of mud exhibited a higher prevalence (37.1%) compared to those from houses constructed with bricks (32.9%). The presence of holes in the wall and the type of windows (closed, open and partial open) were significantly associated with malaria prevalence, whereby participants from households with holes in the walls (39.2%) and open windows (36.3%) had higher prevalence of malaria infections (**Table 3**).

**Table 3.** Prevalence of malaria by household characteristics.

|  | Examined (N) | n (%) mRDT positive | p-values |
| --- | --- | --- | --- |
| Variable |  |  |  |
| Family size |  |  |  |
| <5 people | 2,188 | 656 (30.0) | <0.01 |
| 5 or more people | 7,476 | 2,647 (35.4) |  |
| SES |  |  |  |
| Low | 3,259 | 1,235 (37.9) | <0.01 |
| Moderate | 3,204 | 1,067 (33.3) |  |
| High | 3,201 | 1,001 (31.3) |  |
| Wall type |  |  |  |
| Bricks | 5,794 | 1,908 (32.9) | <0.01 |
| Mud | 3,236 | 1,199 (37.1) |  |
| others | 634 | 196 (30.9) |  |
| Presence of holes in wall |  |  |  |
| Yes | 1,074 | 421 (39.2) | <0.01 |
| No | 8,568 | 2,876 (33.6) |  |
| Windows type |  |  |  |
| Closed | 3,740 | 1,182 (31.6) | <0.01 |
| Open | 4,869 | 1,769 (36.3) |  |
| Partial open | 1,030 | 346 (33.6) |  |

### Factors associated with the risk of asymptomatic malaria infections

Multivariate logistic regression model after adjusting for both individual and household characteristics showed that the odds of malaria infections was higher in males (aOR= 1.32, 95% CI: 1.19 - 1.48, p<0.01) compared to females. In the different age groups, the odds of malaria infection were higher in under-fives (aOR = 2.02, 95% CI: 1.74 - 2.40, p<0.01) and school children ((aged 5 to <10 years old (aOR = 3.23, 95% CI: 2.78 - 3.76, p<0.01) and 10 to <15 years old (aOR = 3.53, 95% CI: 3.03 - 4.11, p<0.01)) compared to adults. Higher odds of malaria infections (aOR 1.49; 95% CI: 1.29 - 1.72, p<0.01) were also observed in individuals who did not sleep under bed nets the night before the survey. Further analysis revealed higher odds of malaria infections in individuals from households with low SES (aOR =1.40, 95% CI: 1.16 - 1.69, p<0.001), and those from houses with open windows (aOR = 1.24, 95% CI: 1.06 - 1.45, p<0.01) and holes in the wall (aOR = 1.43, 95% CI: 1.13 - 1.81, p<0.01) (**Table 4**).

**Table 4.** Socio-demographic and household characteristics associated with the risk of malaria infections among individuals enrolled from five districts.

| **Variables** | **uOR, 95% CI** | **p-value** | **aOR, 95% CI** | **p-value** |
| --- | --- | --- | --- | --- |
| Sex |  |  |  |  |
| Male | 1.57 (1.41 - 1.74) | <0.01 | 1.32 (1.19 - 1.48) | <0.01 |
| Female (reference) | 1 |  | 1 |  |
| Age groups |  |  |  |  |
| <5 years | 2.15 (1.85 - 2.50) | <0.01 | 2.02(1.74 - 2.40) | <0.01 |
| 05 - <10 years | 3.45 (2.97 - 4.00) | <0.01 | 3.23 (2.78 - 3.76) | <0.01 |
| 10 - <15 years | 3.70 (3.18 - 4.29) | <0.01 | 3.53 (3.03 - 4.11) | <0.01 |
| 15+(reference) | 1 |  | 1 |  |
| Use of bed net in the previous night |  |  |  |  |
| Yes (Reference) | 1 |  | 1 |  |
| No | 1.62 (1.41 - 1.86) | <0.01 | 1.49 (1.29 - 1.72) | <0.01 |
| Family size |  |  |  |  |
| <5 people (reference) | 1 |  | 1 |  |
| 5 or more people | 1.28 (1.15 - 1.42) | <0.01 | 1.11 (0.93 - 1.33) | 0.17 |
| SES |  |  |  |  |
| Low | 1.62(1.41 - 1.86) | <0.01 | 1.40 (1.16 - 1.69) | <0.001 |
| Moderate | 1.36 (1.16 - 1.59) | 0.08 | 1.16 (0.97 - 1.40) | 0.11 |
| High (reference) | 1 |  | 1 |  |
| Window types |  |  |  |  |
| Closed (reference) | 1 |  | 1 |  |
| Open | 1.30 (1.13 - 1.51) | <0.01 | 1.24 (1.06 - 1.45) | <0.01 |
| Partial open | 1.12 (0.88 - 1.41) | 0.23 | 1.12 (0.87 - 1.43) | 0.11 |
| Wall type |  |  |  |  |
| Bricks/blocks (reference) | 1 |  | 1 |  |
| Mud | 1.30 (1.12 - 1.50) | <0.01 | 1.10 (0.94 - 1.30) | 0.31 |
| Others | 0.92 (0.69 - 1.23) | 0.57 | 0.83 (0.60 - 1.29) | 0.24 |
| Presence of holes in wall |  |  |  |  |
| Yes | 1.45 (1.17 - 1.81) | <0.01 | 1.43 (1.13 - 1.81) | <0.01 |
| No (reference) | 1 |  | 1 |  |
| Eaves |  |  |  |  |
| Closed (reference) | 1 |  | 1 |  |
| Open | 0.89 (0.78 - 1.02) | 0.1 | 1.09 (0.94 - 1.27) | 0.24 |

CI: Confidence intervals; uOR: Unadjusted odds ratio; aOR: Adjusted odd ratio

The analysis of random effects measures revealed a significant clustering effect of the household with ICC 0.33 indicating that about 33% of total variations in malaria infection can be attributed to household characteristics (null model). The last model with the lowest AIC and likelihood ratio was considered the best model for predicting the association between independent variables and the prevalence of malaria (**Table 5**).

**Table 5.** Random effect model and comparison of the best fit of factors associated with malaria prevalence.

| Parameter | Null model | Model I | Model II |
| --- | --- | --- | --- |
| Cluster level variance (SE) | 1.29(0.15) | 1.88(0.15) | 1.84 (0.14) |
| ICC | 33% | 36% | 36% |
| Fit model criteria |  |  |  |
| LL | -5892.54 | -5628.98 | -5606.18 |
| AIC (-2LL) | 11,785.072 | 11,258 | 11,212.4 |

ICC: Inter-cluster correlation; AIC: Akaike information criterion; LL: Log-likelihoods

## Discussion

Although asymptomatic malaria has been given little attention, there is a necessity to integrate and implement strategies targeting asymptomatic cases in routine malaria control and elimination initiatives due to their potential role as a major reservoir of malaria transmission. Currently, there is limited information on the prevalence and potential drivers associated with asymptomatic infections which would provide evidence for targeting them with specific interventions particularly in the ongoing elimination efforts in Tanzania. This study was conducted to assess the prevalence and drivers of malaria infections among asymptomatic individuals from selected communities in five regions of Tanzania with different transmission intensities. In the current study, the overall prevalence of malaria infections was high (34.4%) and highly variable at the community and regional levels. The study also showed high prevalence and risk of malaria infections among school children, males, participants with fever (as history in the past 2 days or at presentation) and with low SES as well as those living in poorly constructed houses. The high prevalence of malaria reported in this study is comparable to what has been previously reported in Tanzania and elsewhere [29,37,38]. The higher prevalence may be attributed to the presence of factors including vector abundance and breeding sites, lack of proper knowledge on malaria prevention and control measures, and other socioeconomic-related factors resulting in failure to afford proper houses that reduce malaria transmission.

This study reported a higher malaria prevalence in males than females (in all age groups except in a few cases, whereby prevalence was higher in females of some age groups), and this is consistent with other findings reported elsewhere [29,39]. It was also shown that males had higher odds of malaria infection compared to females, possibly due to the lifestyle and activities done by males in rural communities. Males are usually more involved in socio-economic activities such as agriculture, fishing, and grazing in environments that are suitable for mosquito breeding [40,41]. Moreover, males spend most of their time outdoors in social gatherings and cultural events up to peak biting hours compared to females, increasing their exposure to mosquito bites [42,43]. In contrast to females, males are also less likely to seek medical care when they experience malaria-related symptoms, increasing their vulnerability to malaria infections [44–46]. In addition to behavioural and socioeconomic factors, there are sex-specific biological factors, such as post-pubertal hormone changes in males (aged 15+ years), that increase their allure to mosquitoes. Immunity-related factors may also contribute to the increased prevalence of malaria in males [47,48]. Studies have suggested that males experience a delayed clearance of infection compared to females in the absence of treatment [49,50]. As a result, males tend to remain asymptomatic for a longer period, increasing their likelihood of testing positive during surveys.

School children had a higher prevalence above the national average of 21.6% reported elsewhere [51,52]. The odds of being infected with malaria parasites was three times higher among school children compared to adults. The higher odds in this group may be attributed to their involvement in risky activities such as routine night studying sessions, traditional initiation ceremonies, and attending social events occurring outdoors up to late at night, which expose them to mosquito bites [52]. Furthermore, the relatively higher odds of malaria infections among school children may be due to the recent epidemiological shift in the peak burden of asymptomatic malaria infections that previously occurred in under-fives [53,54]. These substantial changes in epidemiological patterns have been attributed to scaled-up interventions such as ITNs, intermittent preventive therapies for pregnant women (IPTp) and infants (IPTi), and improved case management based on prompt diagnosis and treatment [34,55,56]. In this study, children below five years had a higher malaria prevalence and the odds of being infected were two times higher compared to adults. This may be due to the fact that under-fives have not yet developed adequate naturally-acquired immunity to malaria due to the lack of repeated exposure to mosquito bites [57–59]. Hence, control efforts targeting under-fives should continue particularly in areas where malaria transmission is high since the risk is still high.

Among all studied villages in the five districts, three villages (Rubuga, Kitoma and Ruko) in Kyerwa had a relatively higher prevalence (>61.0%) compared to others. Conversely, only four villages had a prevalence of less than 20%, including two (Magoda and Mamboleo) from Muheza district, and one village each from Nyasa (Chiulu) and Kyerwa (Nyakabwera). The prevalence varied significantly across villages, especially in Kyerwa district where villages that are proximate to each other experienced different burdens, suggesting the existence of other potential environmental factors contributing to the micro-geographic pattern of malaria within these communities. For instance, all study villages in Kyerwa (except Nyakabwera) are partly bordered by small lakes which are part of the Kagera river basin, and these provide suitable breeding sites for malaria vectors [60,61]. The variation in malaria burden at the micro-geographic level has been observed in another study conducted in Tanzania [62] that revealed heterogeneity in the risk of malaria in 80 councils. The reasons for low malaria prevalence in villages from other districts could include the efforts done by the government through NMCP to implement effective malaria interventions, including free ITNs distribution campaigns, IPTp and strengthened malaria surveillance systems in areas with high transmission [25,63,64].

The distribution and use of bed nets constitute core interventions for preventing malaria infections in Tanzania [1,2]. The nets offer protection against mosquito bites, effectively reducing the transmission of malaria parasites by mosquitoes and therefore contributing to a significant decrease in malaria risk at both individual and community levels [22,65,66]. In this study, the overall bed net ownership and use on the night before the survey were 78% and 77.2%, respectively. This is relatively higher compared to that reported elsewhere [1,67]. Bed net ownership and use were significantly higher (≥91%) in three districts: Muheza, Ludewa and Nyasa, compared to others. The elevated trend in ownership and bed net use could likely be the results of extensive ITN distribution efforts that have been intensively done by NMCP in the past two decades [68]. In Tanzania, NMCP aimed at ensuring universal access to ITNs at the rate of at least one ITN for every two people and reaching coverage of 80% by 2023 and 85% by 2025 [2]. Additional studies are needed to uncover factors contributing to reduced bed net ownership and usage in Kyerwa and Buhigwe districts.

Bed net ownership and use were higher among females and under-fives across districts and this is not surprising because in Tanzania there is an ongoing campaign focused on providing free bed nets to pregnant women during antenatal care visits. This is to ensure availability of ITNs for protection against malaria for both expectant mothers and their unborn children [64,69]. Under-fives are also given free bed nets during their clinic visits [70]. Likewise, school children have been targeted for controlling and reducing malaria burden in Tanzania for a decade now and a school bed net programme has been running through schools to sustain ITNs access and use [71,72]. Several studies have reported increased bed net ownership among school children in Tanzania [73,74]. Despite the increase in ownership and use rates of bed nets among school children, reaching 75.5% and 75.2%, respectively, this group still exhibited a high prevalence of malaria. Further studies are needed to assess different questions related to bed net ownership and use among school children as well as the reason for the increased malaria burden in this group.

A significantly higher prevalence of malaria infections was observed in families with five or more members, low SES, and those living in houses constructed with mud compared to their counterparts. Furthermore, the presence of holes in the walls and open windows in households was associated with a higher prevalence and risk of malaria infections. These findings align with other studies indicating increased malaria vector biting risk with the increase of household occupants in rural communities [75] and low SES as it decreases the ability to afford malaria prevention, control and treatment services [76– 78]. Additionally, the mud-constructed houses with openings and unscreened windows provide entry points for mosquitoes and consequently result in higher indoor vector densities [66,75]. Conversely, other studies reported a decreasing risk of malaria transmission associated with high-quality housing [79,80]. Therefore, it is critical to enhance awareness of better housing and interventions that reduce malaria transmission. It is also critical to facilitate financial initiatives for improved SES and housing conditions for vector control to expedite malaria elimination efforts.

Individuals who reported not sleeping under bed nets the night before the survey exhibited higher malaria prevalence, in line with what was reported by other studies conducted elsewhere [29,81,82]. The risk of malaria infection in this group was 49% higher than their counterparts. Various studies have shown that sleeping under ITNs offers a physical barrier to mosquito bites and effectively prevents malaria transmission by killing or deterring mosquitoes [65,83]. Therefore, there is a need for educational initiatives to enable rural communities to understand the critical roles bed nets play against malaria transmission [84,85].

The current study found a negative correlation between SES and malaria burden. The risk of malaria among individuals with low SES was 40% higher than those with higher SES, and this is consistent with findings from studies reported elsewhere [59,76,86]. This increased risk could be attributed to various factors, including limited access to healthcare services and preventive measures such as ITNs and IRS [77,87,88]. Most individuals with low SES reside in poorly constructed houses with holes on the wall and unscreened windows associated with increased indoor mosquito bites and significantly increasing their vulnerability to malaria infections [89,90]. Studies have suggested implementing different house modifications to reduce the risk of malaria transmission, including screening windows and doors, repairing walls and using modern roofing materials. Thus, it is critical to devise strategies aimed at addressing SES inequalities [79,91,92] and improving housing conditions in rural communities to accelerate malaria control and eventually elimination.

Limitations to this study include the use of RDTs-based results only, which are less sensitive compared to molecular approaches, particularly polymerase chain reaction (PCR), which would allow to clarify false-negative diagnoses obtained with RDTs [93]. The survey was carried out during the dry season when malaria transmission in some regions is low [94,95], which might have resulted in an underestimation of overall malaria prevalence. The use of convenient sampling may potentially introduce bias since participants may have enrolled due to accessibility, the free services offered, or consultations with project physicians. Since the study population was not selected randomly, it might have varied, possibly resulting in a non-representative sample, which limits the extrapolation of the study findings. However, the results are consistent with previous studies published elsewhere, indicating a low degree of selection bias.

## Conclusion

This study revealed a high prevalence of malaria infections with varying burden at district/regional and village levels. The odds of malaria infection were higher among males, under-five and school children. Individuals with low SES, not using bed nets and living in poorly constructed houses with open windows and holes in the walls had higher odds of malaria infections. The results from this study highlight the need for strong initiatives to control asymptomatic malaria infections and identify vulnerable groups of high priority requiring more intensified control efforts, especially implementing vector control measures such as ITNs and other available interventions to effectively control and eliminate malaria.

## List of abbreviations

ACT: Artemisinin-based combination therapy
AIC: Aike Information content
ANC: Antenatal care clinic
aOR: Adjusted odds ratio
ART-R: Artemisinin partial resistance
CI: Confidence interval
cOR: Crude odds ratios
CSS: Cross-sectional survey
DBS: Dried blood spots
DMFP: District malaria focal person
GPS: Geographic positioning system
ICC: Inter-cluster correlation coefficient
IDs: Identification numbers
IPTp: Intermittent preventive treatment
IQR: Interquartile range
IRS: Indoor residual spraying
ITN: Insecticides treated nets
LSM: Larval source management
MRCC: Medical Research Coordinating Committee
MSMT: Molecular surveillance of malaria in Tanzania.
NIMR: National Institute for Medical Research
NMCP: National Malaria Control Program
ODK: Open Data Kit software
ORs: Odds ratio
PCA: Principal component analysis
PO-RALG: President’s Office, Regional Administration and Local Government
RDTs: Rapid diagnostic tests for malaria
SES: Socio-economic status
SP: Sulfadoxine-pyrimethamine
WHO: World Health Organization
WHO-Afro: WHO Regional office for African

## Declarations

### Ethics approval and consent to participate

This CSS was part of the MSMT projects whose protocol was reviewed and approved by the Medical Research Coordinating Committee (MRCC) of the National Institute for Medical Research (NIMR). Authorization to conduct the study was obtained from the President’s Office, Regional Administration and Local Government (PO-RALG), regional authorities, and the District Executive Directors. Information about the CSS was disseminated in the community through their village mobilisation teams for two consecutive days preceding the survey. Prior to participating in the survey, verbal and written informed consent were sought and obtained from all participants or parents/guardians in case of children.

### Availability of data and materials

The data used in this paper are available and can be obtained upon a request from the corresponding author.

### Authors contribution

DSI developed the idea, supervised study implementation and data analysis, and participated in the interpretation of the results. FF, DPC and DAP were involved in data collection, analysis and results interpretation. DSI and GAC drafted the manuscript and all authors revised and contributed to the final edition. DSI revised and finalised the manuscript and all authors read and approved the manuscript.

## Data Availability

The data used in this paper are available and can be obtained upon request from the corresponding author.

## Acknowledgements

The authors wish to sincerely thank the participants for their willingness to participate in the CSS, providing consent and contributing to the study. They also extend their gratitude to the data collection and laboratory teams for their valuable contributions, including Ezekiel Malecela, Oswald Oscar, Ildephonce Mathias, Gerion Gaudin, Kusa Mchaina, Hussein Semboja, Sharifa Hassan, Salome Simba, Hatibu Athumani, Ambele Lyatinga, Honest Munishi, Anael Derrick Kimaro, Ally Idrissa and Amina Ibrahim. Special thanks to the finance, administrative, and logistic support teams at NIMR: Christopher Masaka, Millen Meena, Beatrice Mwampeta, Neema Manumbu, Arison Ekoni, Sadiki Yusuph, John Fundi, Fred Mashanda, Amir Tununu and Andrew Kimboi. The support from the management of NIMR, NMCP and PO-RALG was critical to the success of this CSS and it is therefore appreciated. The team extends gratitude for the technical and logistics support from partners at Brown University, the University of North Carolina at Chapel Hill, the CDC Foundation and the Bill and Melinda Gates Foundation team.

---

**Supplementary Figure S1:**
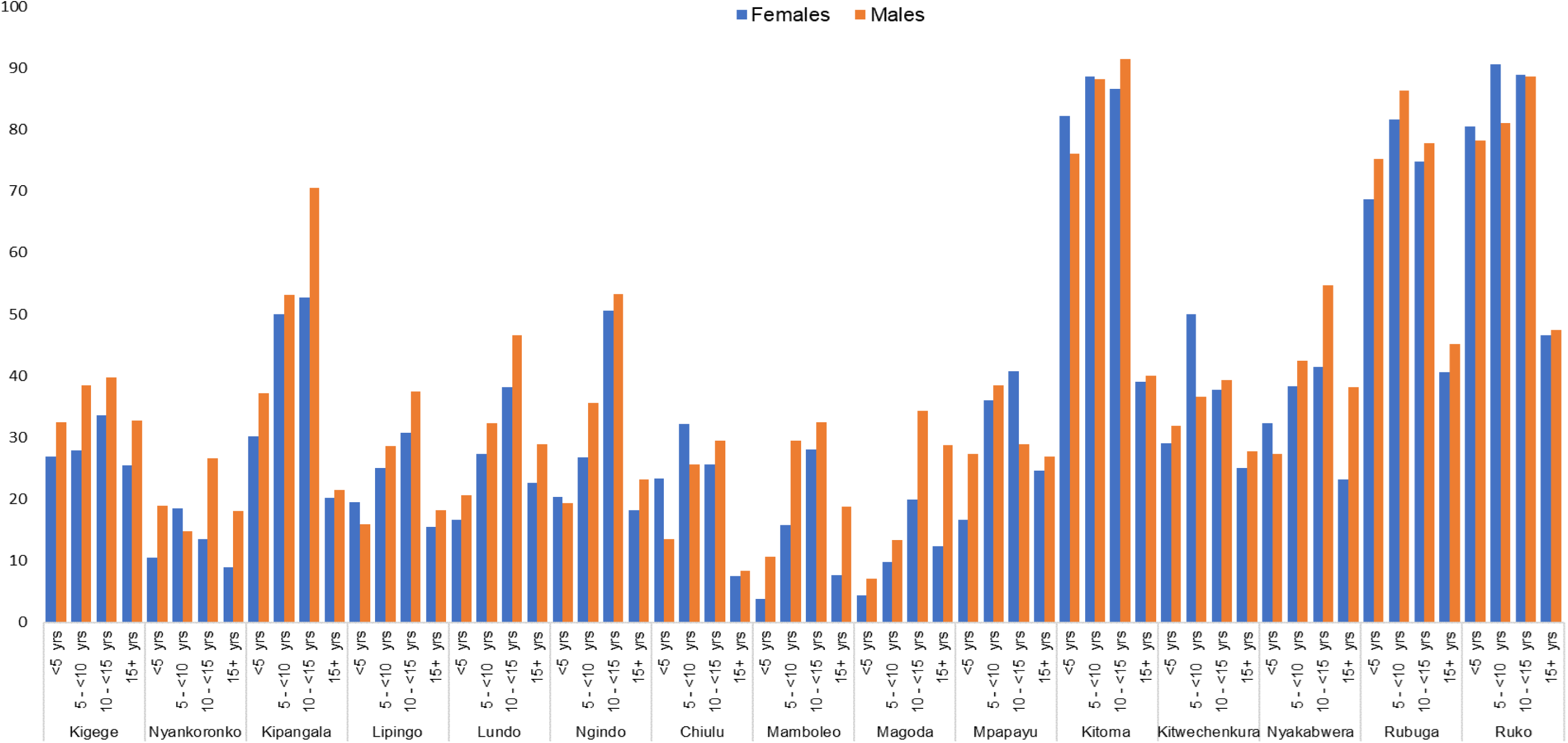
Distribution of malaria prevalence by sex and age groups among participants from different villages in five districts

## References

1. Demographic and Health Survey and Malaria Indicator Survey 2022.

2. NMCP. National Malaria Strategic plan for Tanzania 2021-2025. 2020. Available from: http://api-hidl.afya.go.tz/uploads/library-documents/1641210939-jH9mKCtz.pdf

3. Global-technical-strategy-for-malaria-2016-2030.pdf.

4. Thawer SG, Golumbeanu M, Lazaro S, Chacky F, Munisi K, Aaron S, et al. Spatio-temporal modelling of routine health facility data for malaria risk micro-stratification in mainland Tanzania. Sci Rep. 2023;13:10600.

5. World malaria report 2023. Available from: https://www.who.int/teams/global-malaria-programme/reports/world-malaria-report-2023

6. United States Agency of International De, United States Agency of International Development. Tanzania: Malaria Operational Plan FY 2015. CreateSpace Independent Publishing Platform; 2015.

7. Mitchell CL, Ngasala B, Janko MM, Chacky F, Edwards JK, Pence BW, et al. Evaluating malaria prevalence and land cover across varying transmission intensity in Tanzania using a cross-sectional survey of school-aged children. Malar J. 2022;21:80.

8. Hussein AK, Tarimo D, Reaves EJ, Chacky F, Abade AM, Mwalimu CD, et al. The quality of malaria case management in different transmission settings in Tanzania mainland, 2017-2018. PLOS Glob Public Health. 2023;3:e0002318.

9. Popkin-Hall ZR, Seth MD, Madebe RA, Budodo R, Bakari C, Francis F, et al. Malaria species positivity rates among symptomatic individuals across regions of differing transmission intensities in Mainland Tanzania. J Infect Dis. 2024;229:959–68.

10. Sendor R, Banek K, Kashamuka MM, Mvuama N, Bala JA, Nkalani M, et al. Epidemiology of *Plasmodium malariae* and *Plasmodium ovale spp*. in a highly malaria-endemic country: a longitudinal cohort study in Kinshasa Province, Democratic Republic of Congo. medRxiv. 2023;

11. Odero JO, Nambunga IH, Masalu JP, Mkandawile G, Bwanary H, Hape EE, et al. Genetic markers associated with the widespread insecticide resistance in malaria vector Anopheles funestus populations across Tanzania. Parasit Vectors. 2024;17:230.

12. Tungu P, Kabula B, Nkya T, Machafuko P, Sambu E, Batengana B, et al. Trends of insecticide resistance monitoring in mainland Tanzania, 2004-2020. Malar J. 2023;22:100.

13. Manguin S, Dev V. Towards Malaria Elimination: A Leap Forward. BoD -- Books on Demand; 2018.

14. World Health Organization. Strategy to respond to antimalarial drug resistance in Africa. Strategy to respond to 2022;

15. White NJ. Antimalarial drug resistance. J Clin Invest. 2004;113:1084–92.

16. Ishengoma DS, Mandara CL, Bakari C, Fola AA, Madebe R, Seth MD, et al. Evidence of artemisinin partial resistance in North-Western Tanzania: Clinical and drug resistance markers study. 2024. Available from: 10.2139/ssrn.4726181

17. Juliano JJ, Giesbrecht DJ, Simkin A, Fola AA, Lyimo BM, Pereus D, et al. Country wide surveillance reveals prevalent artemisinin partial resistance mutations with evidence for multiple origins and expansion of sulfadoxine-pyrimethamine resistance mutations in northwest Tanzania. medRxiv. 2023;

18. Rogier E, Battle N, Bakari C, Seth MD, Nace D, Herman C, et al. *Plasmodium falciparum pfhrp2* and *pfhrp3* gene deletions among patients enrolled at 100 health facilities throughout Tanzania: February to July 2021. Sci Rep. 2024;14:8158.

19. Gamboa D, Ho M-F, Bendezu J, Torres K, Chiodini PL, Barnwell JW, et al. A large proportion of P. falciparum isolates in the Amazon region of Peru lack *pfhrp2* and *pfhrp3*: implications for malaria rapid diagnostic tests. PLoS One. 2010;5:e8091.

20. Al-Eryani SM, Irish SR, Carter TE, Lenhart A, Aljasari A, Montoya LF, et al. Public health impact of the spread of *Anopheles stephensi* in the WHO Eastern Mediterranean Region countries in Horn of Africa and Yemen: need for integrated vector surveillance and control. Malar J. 2023;22:187.

21. Tadesse FG, Ashine T, Teka H, Esayas E, Messenger LA, Chali W, et al. *Anopheles stephensi* Mosquitoes as Vectors of *Plasmodium vivax* and *P. falciparum*, Horn of Africa, 2019. Emerg Infect Dis. 2021;27:603–7.

22. Ochomo E, Chahilu M, Cook J, Kinyari T, Bayoh NM, West P, et al. Insecticide-Treated Nets and Protection against Insecticide-Resistant Malaria Vectors in Western Kenya. Emerg Infect Dis. 2017;23:758–64.

23. World Health Organization. Seasonal malaria chemoprevention with sulfadoxine--pyrimethamine plus amodiaquine in children: a field guide. Genève, Switzerland: World Health Organization; 2023.

24. Mushi V, Mbotwa CH, Zacharia A, Ambrose T, Moshi FV. Predictors for the uptake of optimal doses of sulfadoxine-pyrimethamine for intermittent preventive treatment of malaria during pregnancy in Tanzania: further analysis of the data of the 2015-2016 Tanzania demographic and health survey and malaria indicator survey. Malar J. 2021;20:75.

25. Joseph JJ, Mkali HR, Reaves EJ, Mwaipape OS, Mohamed A, Lazaro SN, et al. Improvements in malaria surveillance through the electronic Integrated Disease Surveillance and Response (eIDSR) system in mainland Tanzania, 2013-2021. Malar J. 2022;21:321.

26. Hershey CL, Bhattarai A, Florey LS, McElroy PD, Nielsen CF, Yé Y, et al. Implementing Impact Evaluations of Malaria Control Interventions: Process, Lessons Learned, and Recommendations. Am J Trop Med Hyg. 2017;97:20–31.

27. Ahmad A, Mohammed NI, Joof F, Affara M, Jawara M, Abubakar I, et al. Asymptomatic *Plasmodium falciparum* carriage and clinical disease: a 5-year community-based longitudinal study in The Gambia. Malar J. 2023;22:82.

28. U.S. President’s Malaria Initiative Tanzania (Mainland) Malaria Profile. FY-2024-Tanzania-Mainland-Country-Profile.pdf.

29. Mandai SS, Francis F, Challe DP, Seth MD, Madebe RA, Petro DA, et al. High prevalence and risk of malaria among asymptomatic individuals from villages with high rates of artemisinin partial resistance in Kyerwa district, North-western Tanzania. 2023.

30. Liheluka EA, Massawe IS, Chiduo MG, Mandara CI, Chacky F, Ndekuka L, et al. Community knowledge, attitude, practices and beliefs associated with persistence of malaria transmission in North-western and Southern regions of Tanzania. Malar J. 2023;22:304.

31. Bakari C, Jones S, Subramaniam G, Mandara CI, Chiduo MG, Rumisha S, et al. Community-based surveys for *Plasmodium falciparum pfhrp2* and *pfhrp3* gene deletions in selected regions of mainland Tanzania. Malar J. 2020;19:391.

32. Ishengoma DS, Francis F, Mmbando BP, Lusingu JPA, Magistrado P, Alifrangis M, et al. Accuracy of malaria rapid diagnostic tests in community studies and their impact on treatment of malaria in an area with declining malaria burden in north-eastern Tanzania. Malar J. 2011;10:176.

33. Ishengoma DS, Mmbando BP, Segeja MD, Alifrangis M, Lemnge MM, Bygbjerg IC. Declining burden of malaria over two decades in a rural community of Muheza district, north-eastern Tanzania. Malar J. 2013;12:338.

34. Ishengoma DS, Mmbando BP, Mandara CI, Chiduo MG, Francis F, Timiza W, et al. Trends of *Plasmodium falciparum* prevalence in two communities of Muheza district North-eastern Tanzania: correlation between parasite prevalence, malaria interventions and rainfall in the context of re-emergence of malaria after two decades of progressively declining transmission. Malar J. 2018;17:252.

35. National Guidelines for Malaria Diagnosis, Treatment and Preventive Therapies. 2020. pdf.

36. Standard Treatment Guidelines & National Essential Medicines List Tanzania Mainland. 2017.pdf.

37. Hayuma PM, Wang CW, Liheluka E, Baraka V, Madebe RA, Minja DTR, et al. Prevalence of asymptomatic malaria, submicroscopic parasitaemia and anaemia in Korogwe District, north-eastern Tanzania. Malar J. 2021;20:424.

38. Arikawa S, Tchankoni MK, Gbeasor-Komlanvi FA, Atekpe SP, Atcha-Oubou T, Figueroa-Romero A, et al. Prevalence and risk factors associated with malaria infection in children under two years of age in southern Togo prior to perennial malaria chemoprevention implementation. Malar J. 2023;22:357.

39. Aikambe JN, Mnyone LL. Retrospective Analysis of Malaria Cases in a Potentially High Endemic Area of Morogoro Rural District, Eastern Tanzania. Res Rep Trop Med. 2020;11:37–44.

40. Paul P, Kangalawe RYM, Mboera LEG. Land-use patterns and their implication on malaria transmission in Kilosa District, Tanzania. Trop Dis Travel Med Vaccines. 2018;4:6.

41. Sharma RK, Singh MP, Saha KB, Bharti PK, Jain V, Singh PP, et al. Socio-economic & household risk factors of malaria in tribal areas of Madhya Pradesh, central India. Indian J Med Res. 2015;141:567–75.

42. Moshi IR, Manderson L, Ngowo HS, Mlacha YP, Okumu FO, Mnyone LL. Outdoor malaria transmission risks and social life: a qualitative study in South-Eastern Tanzania. Malar J. 2018;17:397.

43. Rodríguez-Rodríguez D, Katusele M, Auwun A, Marem M, Robinson LJ, Laman M, et al. Human Behavior, Livelihood, and Malaria Transmission in Two Sites of Papua New Guinea. J Infect Dis. 2021;223:S171–86.

44. Ngasala B, Mwaiswelo RO, Chacky F, Molteni F, Mohamed A, Lazaro S, et al. Malaria knowledge, attitude, and practice among communities involved in a seasonal malaria chemoprevention study in Nanyumbu and Masasi districts, Tanzania. Front Public Health. 2023;11:976354.

45. Hasabo EA, Khalid RI, Mustafa GE, Taha RE, Abdalla RS, Mohammed RA, et al. Treatment-seeking behaviour, awareness and preventive practice toward malaria in Abu Ushar, Gezira state, Sudan: a household survey experience from a rural area. Malar J. 2022;21:182.

46. Mburu CM, Bukachi SA, Shilabukha K, Tokpa KH, Ezekiel M, Fokou G, et al. Determinants of treatment-seeking behavior during self-reported febrile illness episodes using the socio-ecological model in Kilombero District, Tanzania. BMC Public Health. 2021;21:1075.

47. Okiring J, Epstein A, Namuganga JF, Kamya EV, Nabende I, Nassali M, et al. Gender difference in the incidence of malaria diagnosed at public health facilities in Uganda. Malar J. 2022;21:22.

48. Kurtis JD, Mtalib R, Onyango FK, Duffy PE. Human resistance to *Plasmodium falciparum* increases during puberty and is predicted by dehydroepiandrosterone sulfate levels. Infect Immun. 2001;69:123–8.

49. Briggs J, Teyssier N, Nankabirwa JI, Rek J, Jagannathan P, Arinaitwe E, et al. Sex-based differences in clearance of chronic *Plasmodium falciparum* infection. Elife. 2020;9. Available from: 10.7554/eLife.59872

50. Rodriguez-Barraquer I, Arinaitwe E, Jagannathan P, Kamya MR, Rosenthal PJ, Rek J, et al. Quantification of anti-parasite and anti-disease immunity to malaria as a function of age and exposure. Elife. 2018;7. Available from: 10.7554/eLife.35832

51. Chacky F, Runge M, Rumisha SF, Machafuko P, Chaki P, Massaga JJ, et al. Nationwide school malaria parasitaemia survey in public primary schools, the United Republic of Tanzania. Malar J. 2018;17:452.

52. Kihwele F, Gavana T, Makungu C, Msuya HM, Mlacha YP, Govella NJ, et al. Exploring activities and behaviours potentially increases school-age children’s vulnerability to malaria infections in south-eastern Tanzania. Malar J. 2023;22:293.

53. Mlugu EM, Minzi O, Kamuhabwa AAR, Aklillu E. Prevalence and Correlates of Asymptomatic Malaria and Anemia on First Antenatal Care Visit among Pregnant Women in Southeast, Tanzania. Int J Environ Res Public Health. 2020;17. Available from: 10.3390/ijerph17093123

54. Yimam Y, Nateghpour M, Mohebali M, Abbaszadeh Afshar MJ. A systematic review and meta-analysis of asymptomatic malaria infection in pregnant women in Sub-Saharan Africa: A challenge for malaria elimination efforts. PLoS One. 2021;16:e0248245.

55. Musoke D, Atusingwize E, Namata C, Ndejjo R, Wanyenze RK, Kamya MR. Integrated malaria prevention in low- and middle-income countries: a systematic review. Malar J. 2023;22:79.

56. Nkumama IN, O’Meara WP, Osier FHA. Changes in Malaria Epidemiology in Africa and New Challenges for Elimination. Trends Parasitol. 2017;33:128–40.

57. Sarfo JO, Amoadu M, Kordorwu PY, Adams AK, Gyan TB, Osman A-G, et al. Malaria amongst children under five in sub-Saharan Africa: a scoping review of prevalence, risk factors and preventive interventions. Eur J Med Res. 2023;28:80.

58. Ranjha R, Singh K, Baharia RK, Mohan M, Anvikar AR, Bharti PK. Age-specific malaria vulnerability and transmission reservoir among children. Global Pediatrics. 2023;6:100085.

59. Chilot D, Mondelaers A, Alem AZ, Asres MS, Yimer MA, Toni AT, et al. Pooled prevalence and risk factors of malaria among children aged 6–59 months in 13 sub-Saharan African countries: A multilevel analysis using recent malaria indicator surveys. PLoS One. 2023;18:e0285265.

60. Nabatanzi M, Ntono V, Kamulegeya J, Kwesiga B, Bulage L, Lubwama B, et al. Malaria outbreak facilitated by increased mosquito breeding sites near houses and cessation of indoor residual spraying, Kole district, Uganda, January-June 2019. BMC Public Health. 2022;22:1898.

61. Getachew D, Balkew M, Tekie H. Anopheles larval species composition and characterization of breeding habitats in two localities in the Ghibe River Basin, southwestern Ethiopia. Malar J. 2020;19:65.

62. Thawer SG, Golumbeanu M, Munisi K, Aaron S, Chacky F, Lazaro S, et al. The use of routine health facility data for micro-stratification of malaria risk in mainland Tanzania. Malar J. 2022;21:345.

63. Madanitsa M, Barsosio HC, Minja DTR, Mtove G, Kavishe RA, Dodd J, et al. Effect of monthly intermittent preventive treatment with dihydroartemisinin–piperaquine with and without azithromycin versus monthly sulfadoxine–pyrimethamine on adverse pregnancy outcomes in Africa: a double-blind randomised, partly placebo-controlled trial. Lancet. 2023;401:1020–36.

64. Koenker H, Worges M, Kamala B, Gitanya P, Chacky F, Lazaro S, et al. Annual distributions of insecticide-treated nets to schoolchildren and other key populations to maintain higher ITN access than with mass campaigns: a modelling study for mainland Tanzania. Malar J. 2022;21:246.

65. Wubishet MK, Berhe G, Adissu A, Tafa MS. Effectiveness of long-lasting insecticidal nets in prevention of malaria among individuals visiting health centres in Ziway-Dugda District, Ethiopia: matched case-control study. Malar J. 2021;20:301.

66. Mosha JF, Lukole E, Charlwood JD, Wright A, Rowland M, Bullock O, et al. Risk factors for malaria infection prevalence and household vector density between mass distribution campaigns of long-lasting insecticidal nets in North-western Tanzania. Malar J. 2020;19:297.

67. Worges M, Kamala B, Yukich J, Chacky F, Lazaro S, Dismas C, et al. Estimation of bed net coverage indicators in Tanzania using mobile phone surveys: a comparison of sampling approaches. Malar J. 2022;21:379.

68. Kramer K, Mandike R, Nathan R, Mohamed A, Lynch M, Brown N, et al. Effectiveness and equity of the Tanzania National Voucher Scheme for mosquito nets over 10 years of implementation. Malar J. 2017;16:255.

69. ’s Malaria Initiative. Protecting Tanzania’s Most Vulnerable from Malaria. PMI. 2018. Available from: https://www.pmi.gov/protecting-tanzanias-most-vulnerable-from-malaria/

70. Health Organization W. WHO recommendations on antenatal care for a positive pregnancy experience: summary: highlights and key messages from the World Health Organization 2018; Available from: https://apps.who.int/iris/bitstream/handle/10665/259947/WHO-RHR-18.02-eng.pdf

71. Desmon S. A Malaria Prevention Success Story: School-Based Net Distribution. Johns Hopkins Center for Communication Programs - Inspiring Healthy Behavior Worldwide. Johns Hopkins Center for Communication Programs; 2018. Available from: https://ccp.jhu.edu/2018/07/09/malaria-prevention-nets-tanzania-ccp/

72. Lalji S, Ngondi JM, Thawer NG, Tembo A, Mandike R, Mohamed A, et al. School Distribution as Keep-Up Strategy to Maintain Universal Coverage of Long-Lasting Insecticidal Nets: Implementation and Results of a Program in Southern Tanzania. Glob Health Sci Pract. 2016;4:251–63.

73. Stuck L, Chacky F, Festo C, Lutambi A, Abdul R, Greer G, et al. Evaluation of long-lasting insecticidal net distribution through schools in Southern Tanzania. Health Policy Plan. 2022;37:243–54.

74. Yukich J, Stuck L, Scates S, Wisniewski J, Chacky F, Festo C, et al. Sustaining LLIN coverage with continuous distribution: the school net programme in Tanzania. Malar J. 2020;19:158.

75. Kaindoa EW, Finda M, Kiplagat J, Mkandawile G, Nyoni A, Coetzee M, et al. Housing gaps, mosquitoes and public viewpoints: a mixed methods assessment of relationships between house characteristics, malaria vector biting risk and community perspectives in rural Tanzania. Malar J. 2018;17:298.

76. Kooko R, Wafula ST, Orishaba P. Socioeconomic determinants of malaria prevalence among under five children in Uganda: Evidence from 2018-19 Uganda Malaria Indicator Survey. J Vector Borne Dis. 2023;60:38–48.

77. Degarege A, Fennie K, Degarege D, Chennupati S, Madhivanan P. Improving socioeconomic status may reduce the burden of malaria in sub Saharan Africa: A systematic review and meta-analysis. PLoS One. 2019;14:e0211205.

78. Were V, Buff AM, Desai M, Kariuki S, Samuels A, Ter Kuile FO, et al. Socioeconomic health inequality in malaria indicators in rural western Kenya: evidence from a household malaria survey on burden and care-seeking behaviour. Malar J. 2018;17:166.

79. Bofu RM, Santos EM, Msugupakulya BJ, Kahamba NF, Swilla JD, Njalambaha R, et al. The needs and opportunities for housing improvement for malaria control in southern Tanzania. Malar J. 2023;22:69.

80. Okumu F, Finda M. Key Characteristics of Residual Malaria Transmission in Two Districts in South-Eastern Tanzania-Implications for Improved Control. J Infect Dis. 2021;223:S143–54.

81. Mwangu LM, Mapuroma R, Ibisomi L. Factors associated with non-use of insecticide-treated bed nets among pregnant women in Zambia. Malar J. 2022;21:290.

82. Lindblade KA, Mwandama D, Mzilahowa T, Steinhardt L, Gimnig J, Shah M, et al. A cohort study of the effectiveness of insecticide-treated bed nets to prevent malaria in an area of moderate pyrethroid resistance, Malawi. Malar J. 2015;14:31.

83. Yang G-G, Kim D, Pham A, Paul CJ. A Meta-Regression Analysis of the Effectiveness of Mosquito Nets for Malaria Control: The Value of Long-Lasting Insecticide Nets. Int J Environ Res Public Health [Internet]. 2018;15. Available from: 10.3390/ijerph15030546

84. Onyinyechi OM, Mohd Nazan AIN, Ismail S. Effectiveness of health education interventions to improve malaria knowledge and insecticide-treated nets usage among populations of sub-Saharan Africa: systematic review and meta-analysis. Front Public Health. 2023;11:1217052.

85. Chokkara R, Avudaiappan S, Anitharani M, Eapen A. School-Based Educational Interventions on Prevention and Control of Malaria-A Systematic Review and Meta-Analysis. Am J Trop Med Hyg. 2022;107:827–32.

86. Anjorin S, Okolie E, Yaya S. Malaria profile and socioeconomic predictors among under-five children: an analysis of 11 sub-Saharan African countries. Malar J. 2023;22:55.

87. Mhango P, Malata MP, Chipeta E, Sixpence A, Taylor TE, Wilson ML, et al. Barriers to accessing malaria treatment amongst school-age children in rural Malawi. Malar J. 2023;22:258.

88. Carrasco-Escobar G, Fornace K, Benmarhnia T. Mapping socioeconomic inequalities in malaria in Sub-Sahara African countries. Sci Rep. 2021;11:15121.

89. Searle KM, Earland D, Francisco A, Muhiro V, Novela A, Ferrão J. Household structure is independently associated with malaria risk in rural Sussundenga, Mozambique. Frontiers in Epidemiology. 2023;3. Available from: https://www.frontiersin.org/articles/10.3389/fepid.2023.1137040

90. Ngadjeu CS, Doumbe-Belisse P, Talipouo A, Djamouko-Djonkam L, Awono-Ambene P, Kekeunou S, et al. Influence of house characteristics on mosquito distribution and malaria transmission in the city of Yaoundé, Cameroon. Malar J. 2020;19:53.

91. Fox T, Furnival-Adams J, Chaplin M, Napier M, Olanga EA. House modifications for preventing malaria. Cochrane Database Syst Rev. 2022;10:CD013398.

92. Yaro JB, Tiono A, Sanou A, Toe HK, Bradley J, Ouedraogo A, et al. Risk factors associated with house entry of malaria vectors in an area of Burkina Faso with high, persistent malaria transmission and high insecticide resistance. Research Square. Research Square; 2021. Available from: 10.21203/rs.3.rs-273831/v1

93. Ngasala B, Chacky F, Mohamed A, Molteni F, Nyinondi S, Kabula B, et al. Evaluation of Malaria Rapid Diagnostic Test Performance and pfhrp2 deletion in Tanzania School Surveys, 2017. Am J Trop Med Hyg. 2024;110:887–91.

94. Tarimo BB, Nyasembe VO, Ngasala B, Basham C, Rutagi IJ, Muller M, et al. Seasonality and transmissibility of *Plasmodium ovale* in Bagamoyo District, Tanzania. Parasit Vectors. 2022;15:56.

95. Duque C, Lubinda M, Matoba J, Sing’anga C, Stevenson J, Shields T, et al. Impact of aerial humidity on seasonal malaria: an ecological study in Zambia. Malar J. 2022;21:325.

